# Does the Wim Hof Method have a beneficial impact on physiological and psychological outcomes in healthy and non-healthy participants? A systematic review

**DOI:** 10.1101/2023.05.28.23290653

**Authors:** Omar Almahayni, Lucy Hammond

## Abstract

**Introduction:** Wim Hof, also known as the iceman, developed a method called Wim Hof Method which he claims to have several benefits on physical and mental health. The aim of this systematic review is to identify and synthesise the results of the studies conducted on Wim Hof Method on physiological and psychological health-related outcomes.

**Materials and Methods:** Medline and Web of Science were searched. Studies were included if they met the predetermined inclusion/exclusion criteria. Data extraction and quality assessment were performed on the included studies. The effects of Wim Hof Method were categorised into physiological or psychological related outcomes and narrative synthesis was conducted.

**Results:** Nine papers were included in this review which consisted of eight individual trials. The findings of this systematic review suggest that the Wim Hof Method may affect the reduction of inflammation in healthy and non-healthy participants as it increases epinephrine levels, causing an increase in interleukin-10 and a decrease in pro-inflammatory cytokines. Additionally, Wim Hof breathing method was suggested to not enhance the performance of an exercise as minute ventilation, tidal volume, and breathing frequency were statistically insignificant.

**Conclusion:** Taken together, the findings of this review show promising use of Wim Hof Method in the inflammatory response category. The focus of future studies should move away from investigating the use of Wim Hof breathing method to enhance exercise performance and towards exploring the benefits of Wim Hof Method in non-healthy participants with inflammatory disorders.

## Introduction

Wim Hof, also known as the iceman, is recognized for his ability to resist extreme cold temperatures. He holds 21 Guinness World Records for some of his remarkable human endurance achievements, including climbing Mount Kilimanjaro while wearing shorts, swimming 66 metres beneath ice, standing for two hours in a container filled with ice cubes, and running a half marathon over the Arctic Circle, only wearing shorts and barefoot [1]. He credits his achievements to the Wim Hof Method (WHM) which consists of three different pillars: Wim Hof breathing method (WHBM), cold therapy, and commitment. According to Wim Hof, a combination of these three pillars enhances a person’s autonomic nervous and immune systems, thus strengthening a person’s health and mentality. WHBM consists of hyperventilation 30-40 times then holding the breath voluntarily at low lung volume [2]. Cold therapy considers taking daily cold showers or sitting in an ice bath [3]. The final pillar, which is commitment, is the foundation of the other two pillars; mastering both mindful breathing and cold exposure takes patience and persistence [4]. To support his claims, Wim Hof has submitted his method and himself for research. Many studies, including case studies, randomised control trials (RCTs), and observational trials have been conducted on him and his method [5, 6, 7, 8, 9, 10, 11, 12, 13, 14, 15, 16].

To identify some of the benefits people are experiencing, Wim Hof collaborated with RMIT University by conducting a survey inquiring about the impact WHM had on an individual’s health and well-being [17]. Over 3,200 people answered the survey, and the majority of the answers were positive. Respondents reported a good mix of physical and mental benefits such as an increase in their tolerance to resist cold, as well as an increase in energy, mood, mental focus, and general health. The survey findings also claimed that the WHM had benefits for specific conditions such as stress, tiredness and fatigue, anxiety, depression, back pain, insomnia, arthritis, and chronic pain [17].

Collectively, these findings appear to suggest promising evidence that the WHM may be of importance to healthy and non-healthy individuals and that the WHM approach may be given consideration in the public health and lifestyle medicine fields. The notion of lifestyle medicine refers to the study of how actions and habits affect illness prevention and treatment [18]. However, caution should be applied insofar as the aforementioned survey [17] has not been peer-reviewed or published, and Wim Hof’s own involvement in this study and others [5, 9] may give rise to conflicts of interest in the pursuit of both an evidence base and a commercial opportunity. Wim Hof has attained a level of celebrity as a result of his achievements, method and associated media career. Due to all his remarkable achievements, his charisma, and authenticity, Wim Hof might be considered to be both micro and achieved celebrity [19], where achieved celebrity refers to the attainment of fame based on recognized talent, achievement, or ability and micro celebrity describes an individual that achieves status and social media presence by self-broadcasting about niche subjects to a small community of followers. Wim Hof is an example of a celebrity that embodies the para-social relationship, exchanging the allure of intimacy, integrity, and authenticity. The growing number of celebrities giving lifestyle and health advice and scientific knowledge to the public indicates the need to critically examine such advice [20]. Nunan *et al.,* 2021 urge caution for the potential of “the unintended consequences of uncritical endorsement and application of lifestyle medicine including the infiltration of pseudoscience, profiteering, and the potential for widening health inequalities by a continued focus on the ‘individual’” (p229) [21]. Therefore, an independent, systematic synthesis of this evidence is warranted to evaluate the positive health claims of the WHM.

A systematic review has not yet been conducted on the WMH body of evidence. Therefore, an independent, systematic synthesis of this evidence is warranted to evaluate the positive health claims of the WHM. Hence, this review aims to identify and synthesise the results of the studies conducted on WHM on physiological and psychological health-related outcomes.

## Materials and Methods

This systematic review followed the PRISMA guidelines for systematic reviews [22]. The protocol was registered in the International Prospective Register of Systematic Reviews (PROSPERO), registration number CRD42022333209.

### Literature search

MEDLINE and Web of Science databases were used to search for published studies for potential inclusion in this systematic review. The following search phrases were used in combination: Wim Hof; breathing exercise OR breathing technique; meditation OR commitment; cold temperature OR cold exposure OR ice bath OR cold shower Table 1. The filters English language, year=“2014 - July 4, 2022”, and journal articles or observational studies or randomized controlled trials (RCTs) were applied. A filter for the year was applied because the first RCT study performed on WHM was in 2014 [10].

**Table 1.** Search strategy of Medline and Web of Science.

| Database | Search strategy |
| --- | --- |
| Medline | 1. Wim Hof<br>2. Breathing exercise OR Breathing technique<br>3. 1 OR 2<br>4. Meditation OR Commitment<br>5. Cold temperature OR Cold exposure<br>OR Ice bath OR Cold shower<br>6. 3 AND 4 AND 5<br>7. Limit 1 and 6 to (English Language<br>and Year = "2014 – July 4, 2022" and<br>Journal Article or Observational |
|  | Study or Randomized Controlled Trial) |
| Web of Science | 1. Wim Hof<br>2. Breathing exercise OR Breathing technique<br>3. 1 OR 2<br>4. Meditation OR Commitment<br>5. Cold temperature OR Cold exposure<br>OR Ice bath OR Cold shower<br>6. 3 AND 4 AND 5<br>7. Limit 1 and 6 to (English Language and Year = "2014 – July 4, 2022" and Articles)<br>Note: all searches are all fields |

### Study selection

Duplicates were deleted before abstracts were screened. The screening of titles and abstracts was conducted by two independent reviewers. Afterwards, the full texts were screened by the two reviewers independently. Any disagreements were resolved by consensus. Screening was performed on Covidence. Studies were included according to the predetermined inclusion/exclusion criteria. Inclusion criteria included RCTs and cohort studies published in peer-reviewed journals, studies conducted on healthy individuals and people with pre-existing medical conditions (adolescents and adults over the age of 14), studies that included all three pillars (breathing, cold exposure, and commitment) of the WHM as defined above, and studies that only focused on WHBM. Articles just covering WHBM were permitted as it has been deemed more important and studied more frequently than other pillars. Furthermore, exclusion criteria included studies that discussed WHM but are not original experimental research or are not peer-reviewed, studies that included children under the age of 14, and studies that used methods similar to WHM, but not actually WHM, such as tummo meditation. Both methods allow for the controlment of body temperature, and both have similar breathing techniques. Tummo meditation consists of breathing and visualisation techniques which are intended to summon spiritual insight, whereas WHM is not religious and is acquired by exposing oneself to difficult natural circumstances [23]. After deciding on the included studies, the reference lists were hand-searched for other relevant articles.

Data extraction comprised the name of the study, place where the study was conducted, research design, participant demographics, study context, description of intervention and control, and key findings. One reviewer completed the extraction while a second reviewer checked it for accuracy. Furthermore, study quality was assessed using the RoB 2.0 risk bias tool [24] for RCTs and the Scottish Intercollegiate Guidelines Network (SIGN) risk bias tool [25] for cohort studies. Two reviewers independently evaluated the possibility of bias. The answers were then compared, and any disagreement was discussed between the reviewers.

### Data synthesis

A narrative synthesis of the findings from the studies was included by providing a description and comparison between the different studies. This comprised a discussion of the contexts in which the studies have been undertaken, and the effects of WHM thematically grouped into physiological or psychological related outcomes. In each thematic grouping, different sets of outcomes were included. The data were not suitable for meta-analysis as there were various unrelated outcome measures resulting in heterogeneity across studies.

## Results

### Study characteristics

Nine papers, seen in Fig 1, published between 2014 and July 4, 2022, were identified that examined the WHM. One of these articles was identified from hand-searching the included studies’ reference lists [6]. Two of these articles [13, 16] were further analyses of a study [10]. Table 2 summarises the characteristics of the listed studies. Of the nine papers included, there were eight individual trials. Four of the trials performed the full WHM [7, 10, 12, 15] and four did only WHBM [6, 8, 11, 15]. The application of WHBM differed between studies as some studies had one breathing exercise [6, 8, 11, 12] while others had two breathing exercises [7, 10, 13, 15, 16]. The first breathing exercise was common in all studies. Cold exposure was conducted according to what is accessible to each study. However, all studies agreed on daily exposure to the cold through having cold showers, but the duration of exposure under the shower differed across studies. The meditation technique was the least focused component of the WHM as it is just a foundation for the other two pillars. All studies had the same description of the meditation technique except for Petraskova Touskova *et al.* (2022). All the authors described it as visualisations meant to promote complete relaxation, while Petraskova Touskova *et al.,* 2022 described it as a form of focusing to obtain self-awareness and will-power. Out of the nine papers, five conducted a randomised controlled trial (RCT) design [10, 11, 13, 15, 16], three conducted a crossover RCT design [6, 7, 8], and one conducted a prospective cohort design [12].

**Figure 1.**
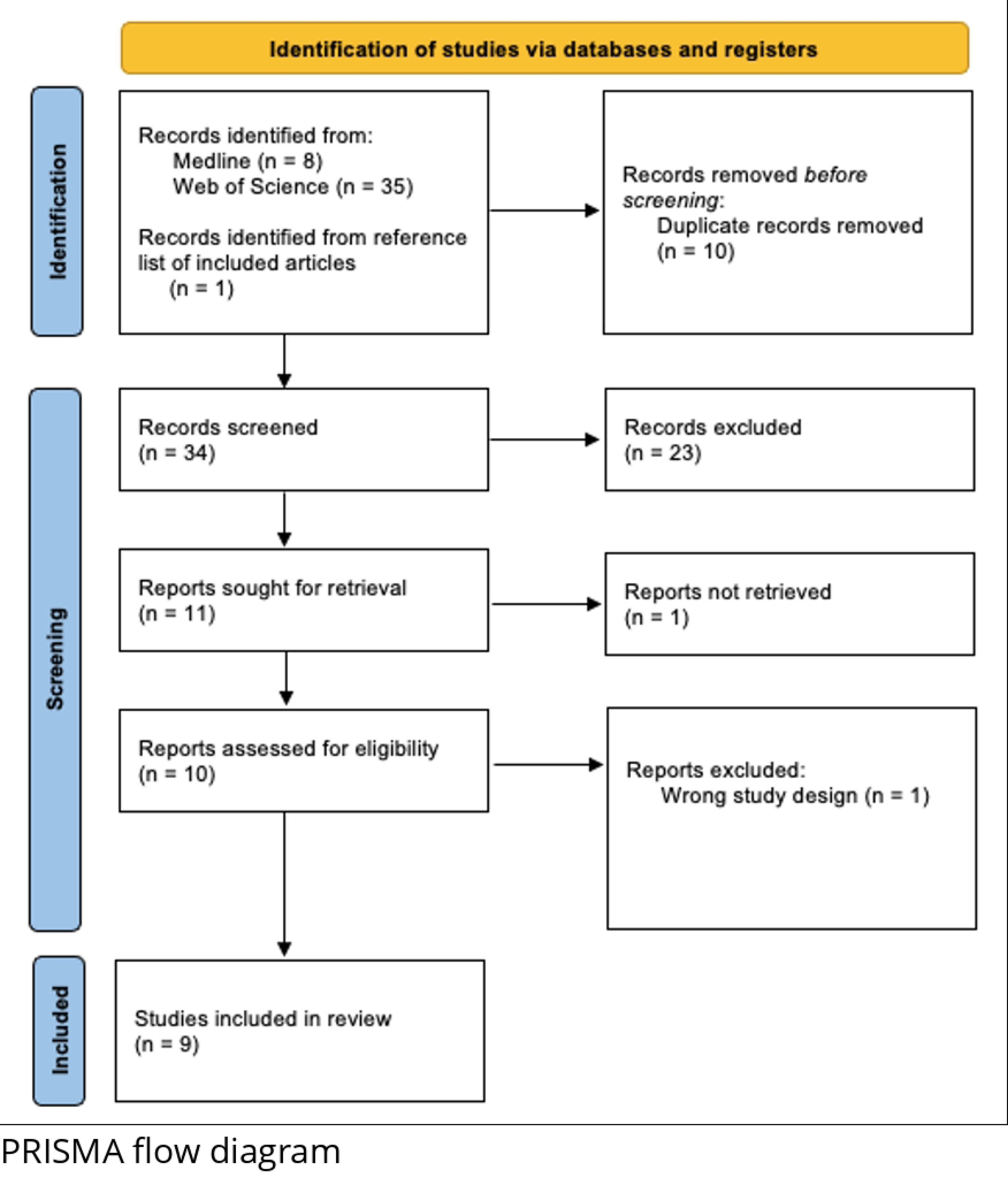
PRISMA flow diagram for study selection processes.

**Table 2.** Characteristics of the listed studies.

| Study | Country | Study Design | Population Description | Endotoxemia Intervention | WHM Intervention | Other Intervention |
| --- | --- | --- | --- | --- | --- | --- |
| Bahenský *et al.*, 2020 | Czech Republic | Crossover RCT | - n=16<br>- Sex: males and females.<br>- Age range: 16-19 years.<br>- Trained: medium and long-distance runners<br>- Running six times every week for at least three years. | N/A | **Intervention**<br>- Breathing exercises were conducted while lying down.<br>**Control**<br>- Rest quietly while sitting down. | **Intervention and control**<br>- Incremental testing session: warm-up (2 min at 25 W), followed by four two-minute stages of increasing intensity (1, 2, 3, 4 W/kg) based on the body mass of each participant. |
| Kox *et al.*,<br>2014 | Netherlands | RCT | - n=24<br>- 30 were recruited, but only 24 completed the full protocol.<br>- Sex: male.<br>- Age range: 19-27 years.<br>- Status: healthy (normal electrocardiography, physical examination, and routine laboratory values). Non-smoking. | **Intervention and control**<br>- iv administration of 2 ng/kg Escherichia coli endotoxin. | **Intervention**<br>- The study was performed in two identical blocks, consisting of nine participants in the WHM-trained group in each block.<br>- Six participated in the endotoxemia experiments and six subjects in the control group.<br>- The WHM-trained group was trained by three trainers and Wim Hof. | N/A |
|  |  |  |  |  | - The day before the endotoxemia experiment, Wim Hof held a last training session in which the chosen participants practised WHM.<br>**Control**<br>- No WHM training. |  |
| Zwaag *et al.*, 2020 | Netherlands | RCT | - Same as Kox *et al.*, 2014. | - Same as Kox *et al.*, 2014. | - Same as Kox *et al.*, 2014. | N/A |
| Middendorp *et al.*, 2016 | Netherlands | RCT | Same as Kox *et al.*, 2014 | - Same as Kox *et al.*, 2014. | - Same as Kox *et al.*, 2014. | N/A |
| Buijze *et al.*,<br>2019 | Netherlands | Open label<br>randomised<br>one-way<br>cross over<br>design | - n=19<br>- 24 were recruited, but only 19 completed the full protocol.<br>- Sex: males and females.<br>- Age range: 18-55 years.<br>- Medical status: clinical diagnosis of axSpA.<br>- Medication use: NSAIDs, corticosteroids, or DMARDs. | N/A | **Early intervention**<br>- Participants were trained by four trainers and Wim Hof.<br>**Late intervention**<br>- The participants in the control group also received exposure to the intervention after the 8-week control phase had ended.<br>- Patients were trained in the same way as the early intervention. | N/A |
|  |  |  |  |  | **Control**<br>- 8 weeks of control period during the early intervention.<br>- No WHM training. |  |
| Zwaag *et al.*, 2022 | Netherlands | RCT | - n=40 for breathing exercises study.<br>- n=48 for endotoxemia study.<br>- Sex: males. | **Intervention and control**<br>- Purified LPS was reconstituted in 5 mL of saline 0.9% for injection, vortexed for 20 | **Endotoxemia study**<br>**Intervention**<br>- Cold exposure alone (CEX).<br>- Breathing exercise without retention alone (BRT). | **Breathing exercises study**<br>**Intervention**<br>- Extensive or short training by Wim Hof. |
|  |  |  | - Age range: 19-24 years for breathing exercises study.<br>- Age range: 20-24 years for endotoxemia study.<br>- Medical status: healthy. | minutes, and then<br>infused as an iv<br>bolus at a dose of<br>2 ng/kg body<br>weight for 1<br>minute at time t =<br>0 hours at<br>9:30AM. | - Cold exposure and the breathing exercise without retention (CBR).<br>**Control**<br>- no WHM training. | - Extensive or short training by an independent trainer.<br>**Control**<br>- No control group. |
| Petraskova<br>Touskova et al., 2022 | Czech<br>Republic | Prospective<br>observational<br>study | - n=13<br>- Sex: females and males.<br>- Age range: 27-60 years. | N/A | **Intervention**<br>- Practised WHM.<br>**Control** | N/A |
|  |  |  | - Status: all had American Society of Anesthesiology health scale scores of 1 to 2.<br>- One male participant was a mild smoker.<br>- The other participants were non-smokers. |  | - No WHM training. |  |
| Citherlet *et al.*, 2021 | Switzerland | Randomised, controlled three-way crossover trial | - n=15<br>- 16 participants enrolled, but 1 was excluded during the trial. | N/A | **Intervention**<br>- Performed WHBM.<br>**Control** | **Intervention**<br>- Hyperventilation.<br>**Control** |
|  |  |  | - Sex: male.<br>- Age range: $\geq 18$ years.<br>- Status: healthy regular runner. |  | - Spontaneous breathing for 12 min. | - N/A |
| Marko *et al.*,<br>2022 | Czech<br>Republic | RCT | - n=19<br>- Sex: males and females.<br>- Age range: 14-19 years.<br>- Status: healthy (free from known metabolic, | N/A | **Intervention**<br>- Performed WHBM for 17-22 mins.<br>**Control**<br>- Breathe naturally while in supine position. | **Intervention**<br>- The test and follow-up test involved a breath-by-breath metabolic study after a graded exercise test (GXT) on a bicycle ergometer. |
|  |  |  | cardiovascular, and/or renal disease).<br>- Trained: middle and long-distance runners. |  |  | - Participants finished four, two-minute stages at 1, 2, 3, and 4 W/kg after a preliminary warm-up at 25 W and a cadence of 80 rev/min.<br>- Throughout the test, participants were told to keep their cadence at around 100 revs/min.<br>- A three-minute relaxation with a cadence of 60 rev/min<br>and resistance of 25 W took place after the final workload phase (4 W/kg).<br>**Control**<br>- N/A |

### Study quality

For the RCT studies, the RoB 2.0 overall risk of bias judgement was ‘high concerns’ for all studies except one study which was judged as ‘some concerns’ [8]. Moreover, the cohort study, using the SIGN tool, was scored ‘unacceptable’ as it was not able to reduce the possibility of bias or confounding.

### Outcomes

Table 3 summarises the WHM description, key findings, and quality assessment. The outcomes were separated into physiological or psychological outcomes. The physiological outcome has six categories: stress response, pro-inflammatory/anti-inflammatory response, metabolites response, respiratory parameters, blood gas measurements, and reporting of symptoms, while psychological outcome has one category: psychological response. The stress response included outcomes such as epinephrine, norepinephrine, dopamine, cortisol, melatonin, and heart rate (HR) whereas pro-inflammatory/anti-inflammatory response included tumour necrosis factor (TNF-α), interleukin-6, interleukin-8, interleukin-10, erythrocyte sedimentation rate (ESR), Ankylosing Spondylitis Disease Activity Score C-reactive protein (ASDAS-CRP), calprotectin, high-sensitivity C-reactive protein (hs-CRP), and Bath Ankylosing Spondylitis Disease Activity Index (BASDAI). Additionally, metabolites response contained lactate and pyruvate while respiratory parameters looked at minute ventilation (V_E_), tidal volume (V_T_), and breathing frequency (BF). The blood gas measurements category included carbon dioxide partial pressure (pCO_2_), pH, and oxygen saturation outcomes and the psychological response category contained expectancy, optimism, neuroticism, short-form 36 (SF-36), EQ-5D, Borg Rating of Perceived Exertion (RPE) scale, and customised questionnaire. Finally, the reporting of symptoms investigated flu-like, self-reported, and depressive symptoms, Trauma Symptom Checklist-40 (TSC-40), and adverse events (AEs).

**Table 3.**
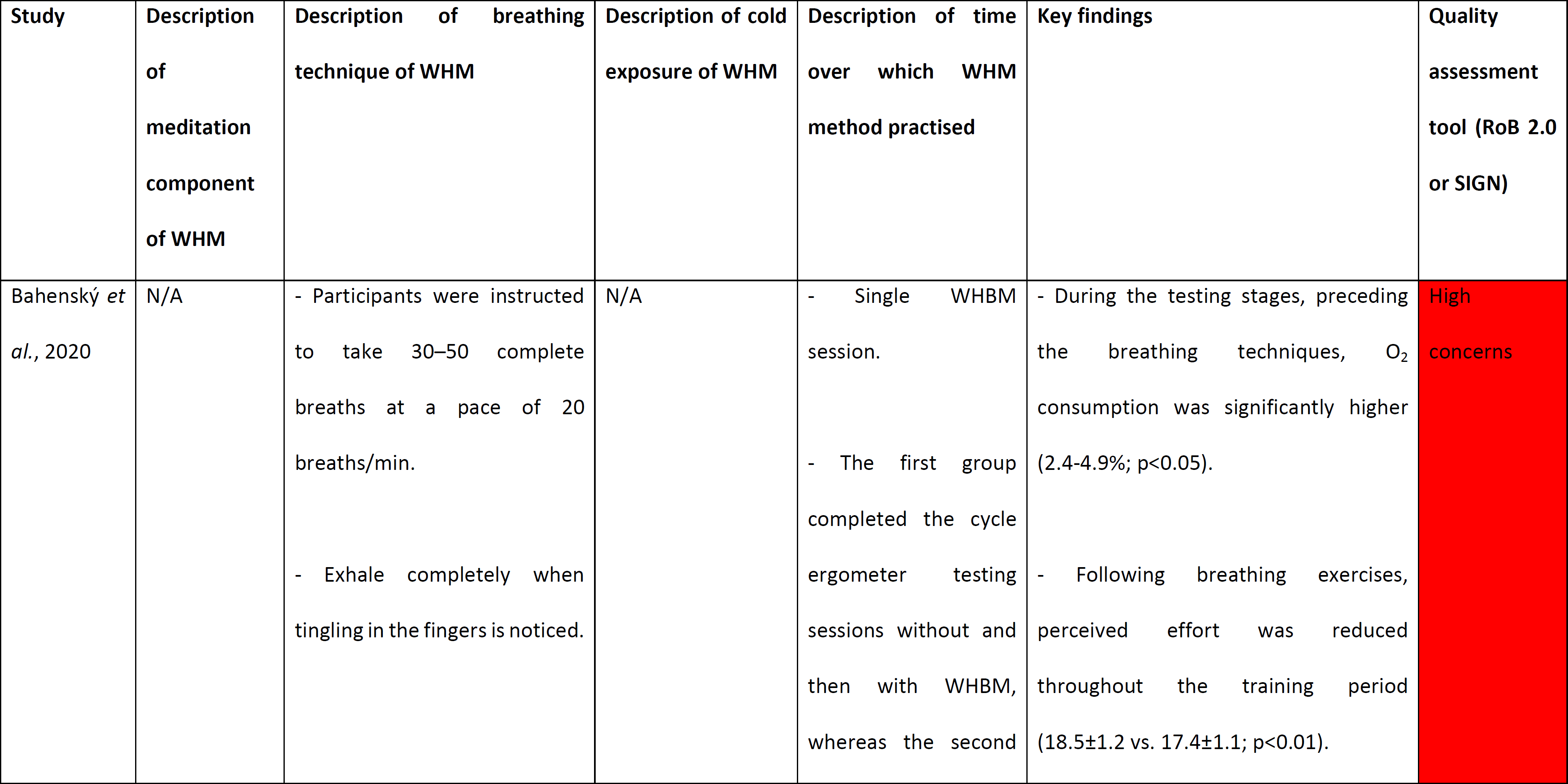
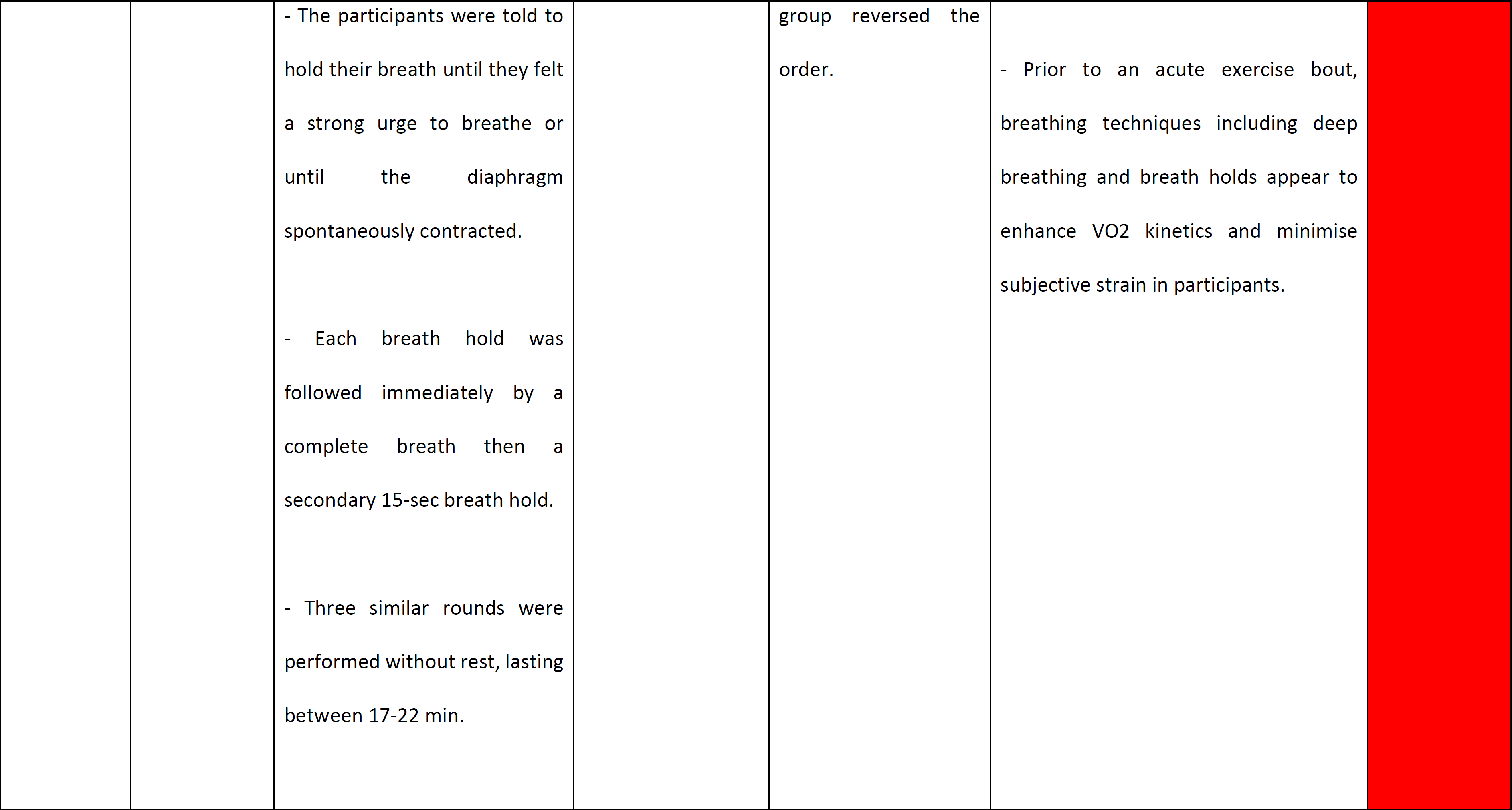
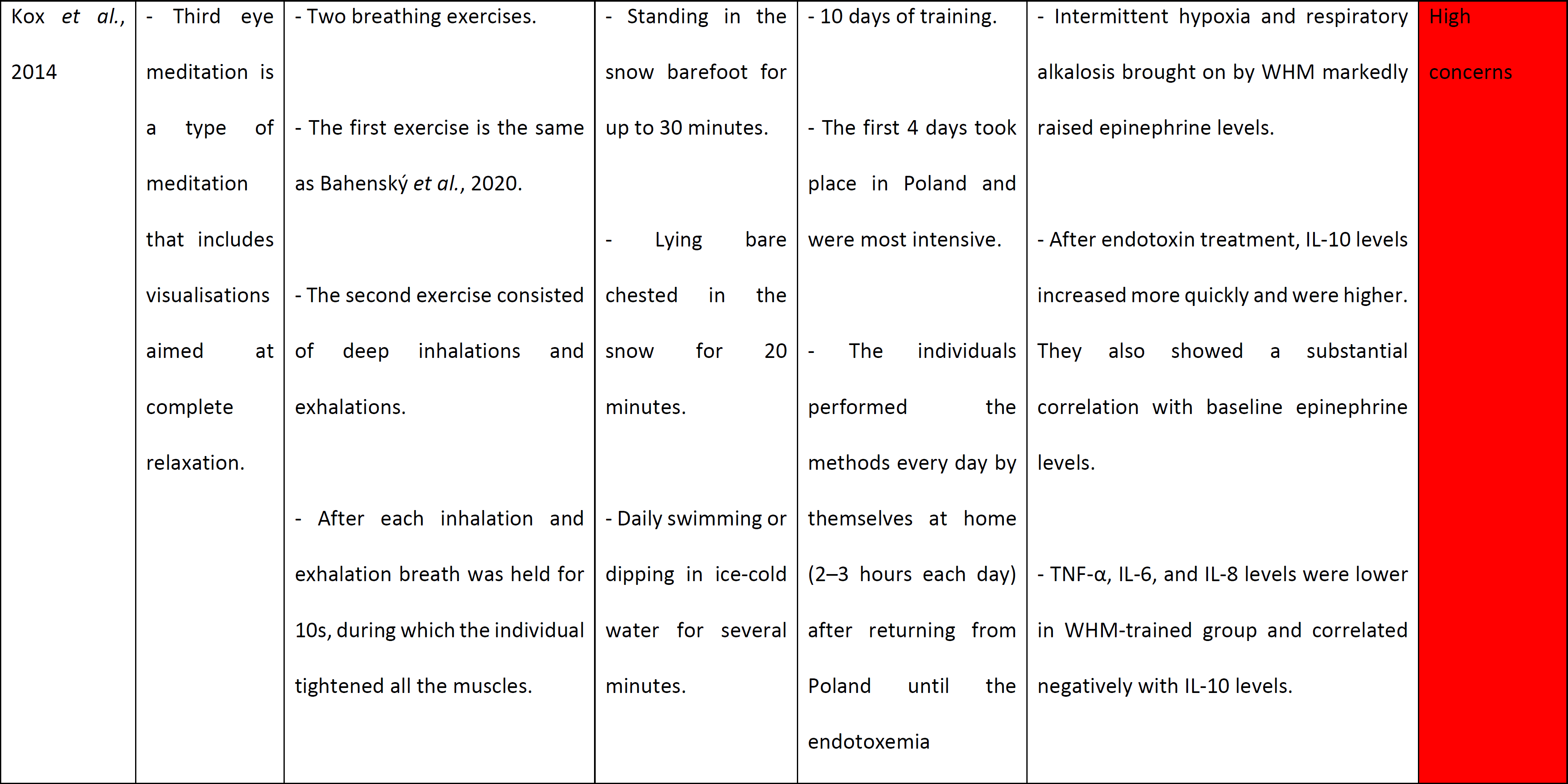
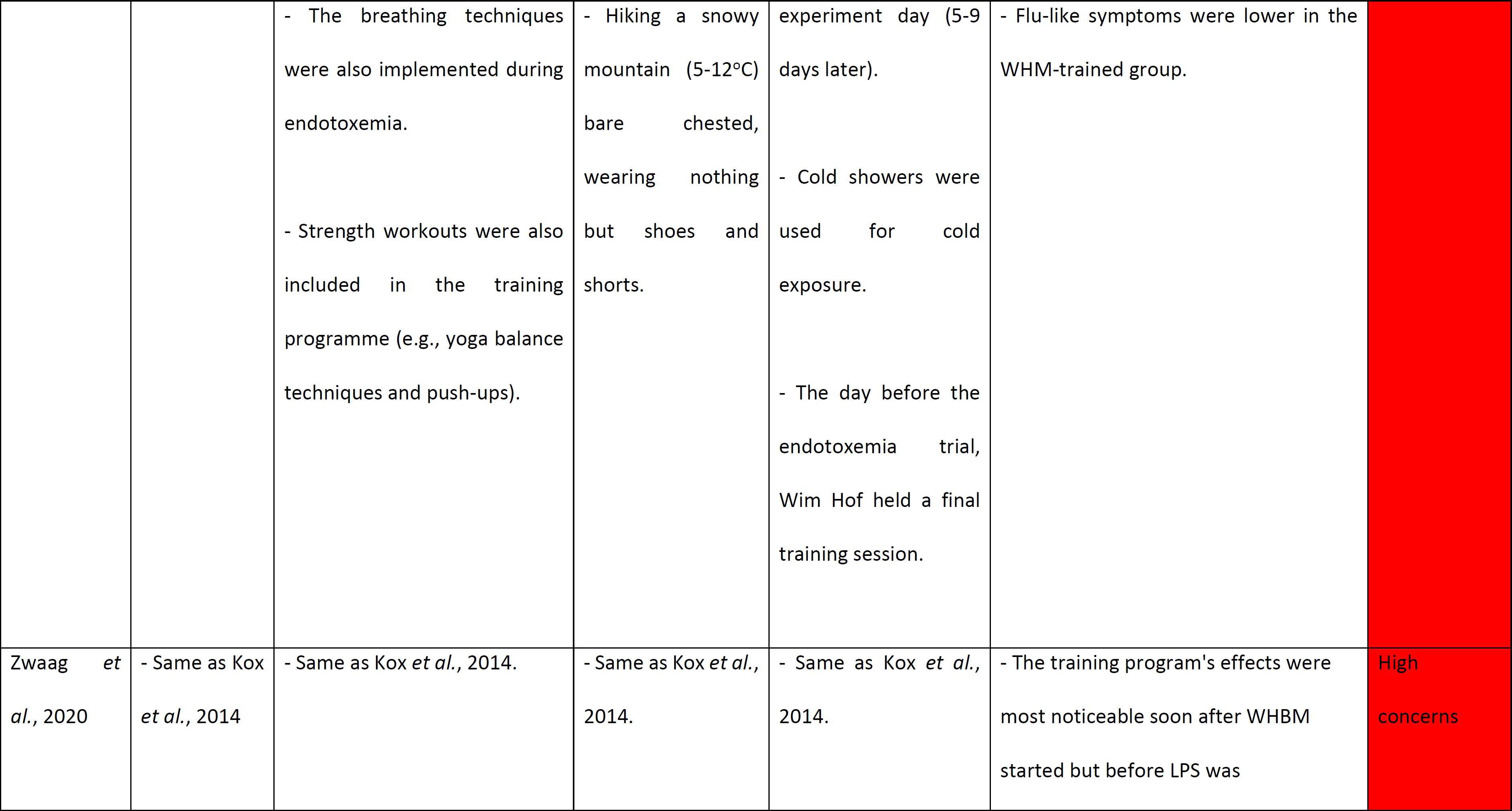
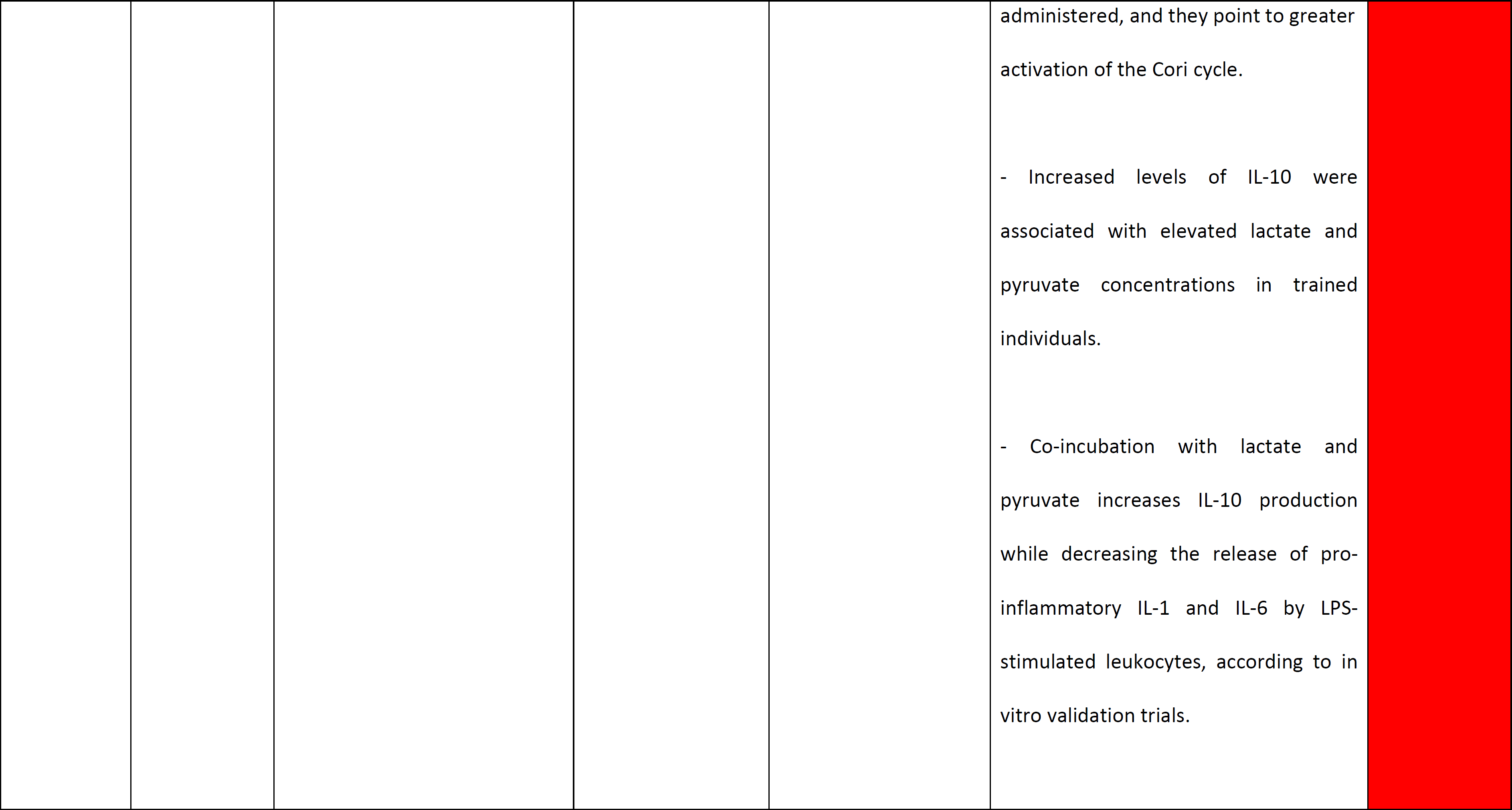
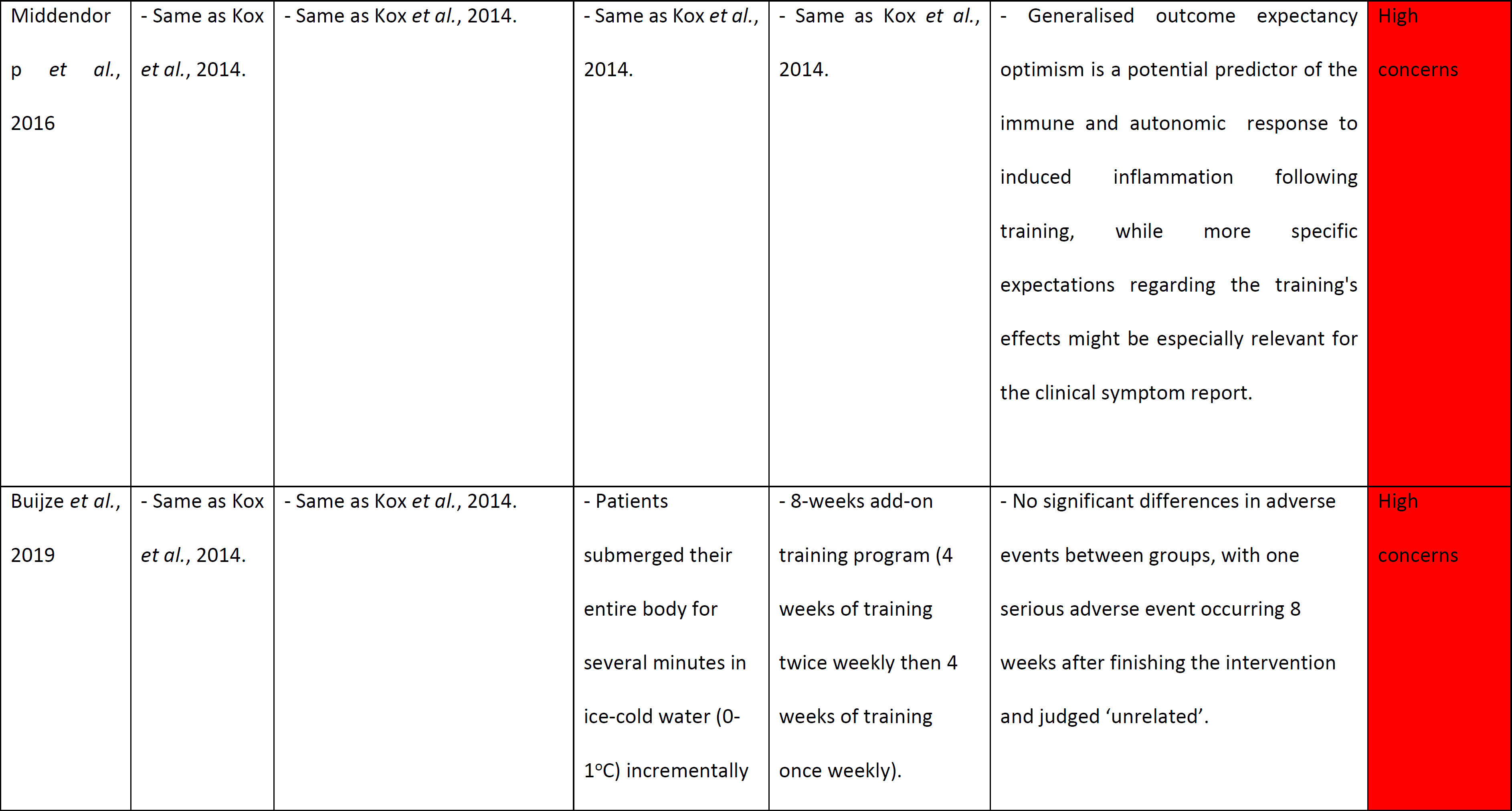
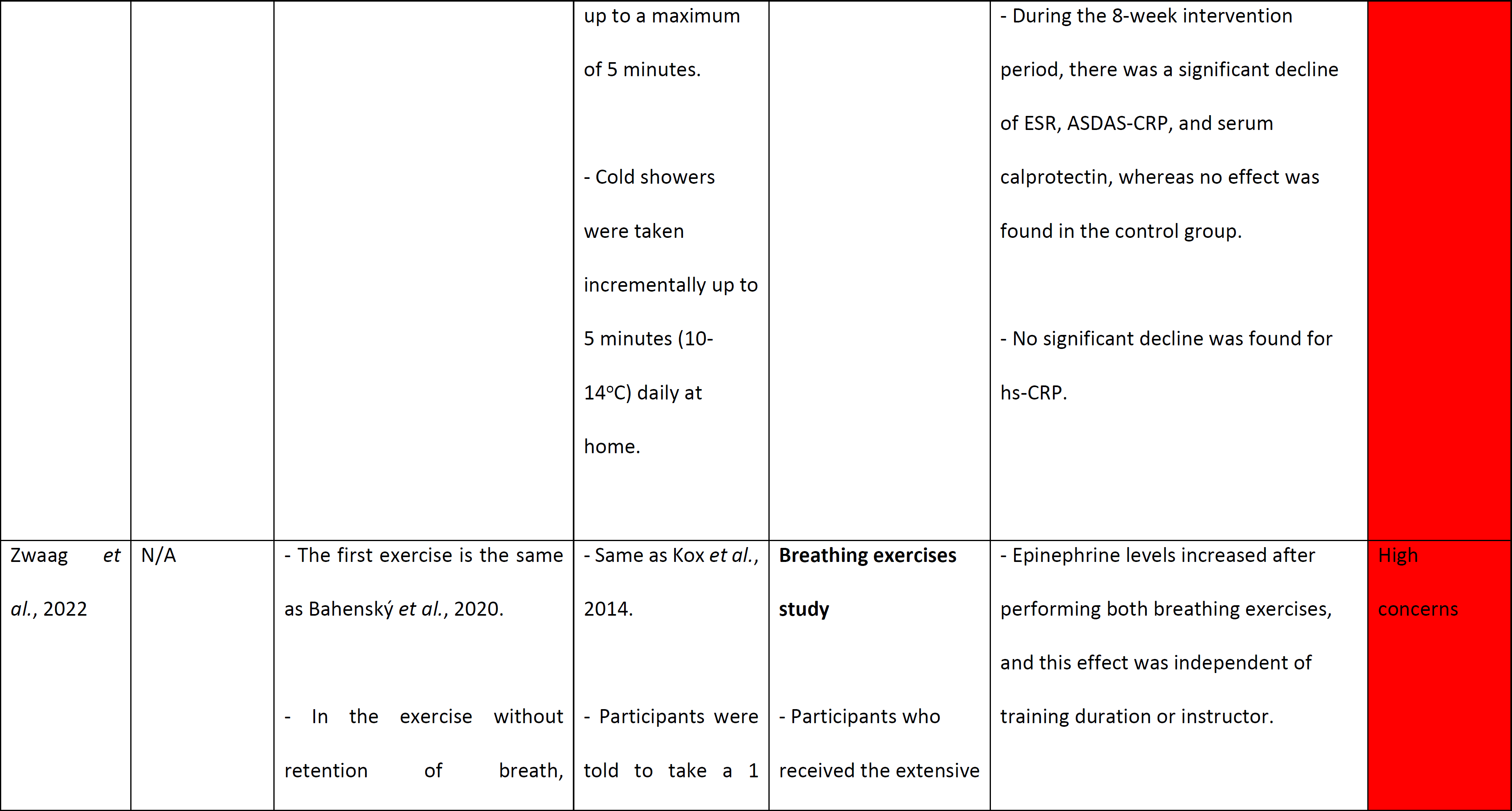
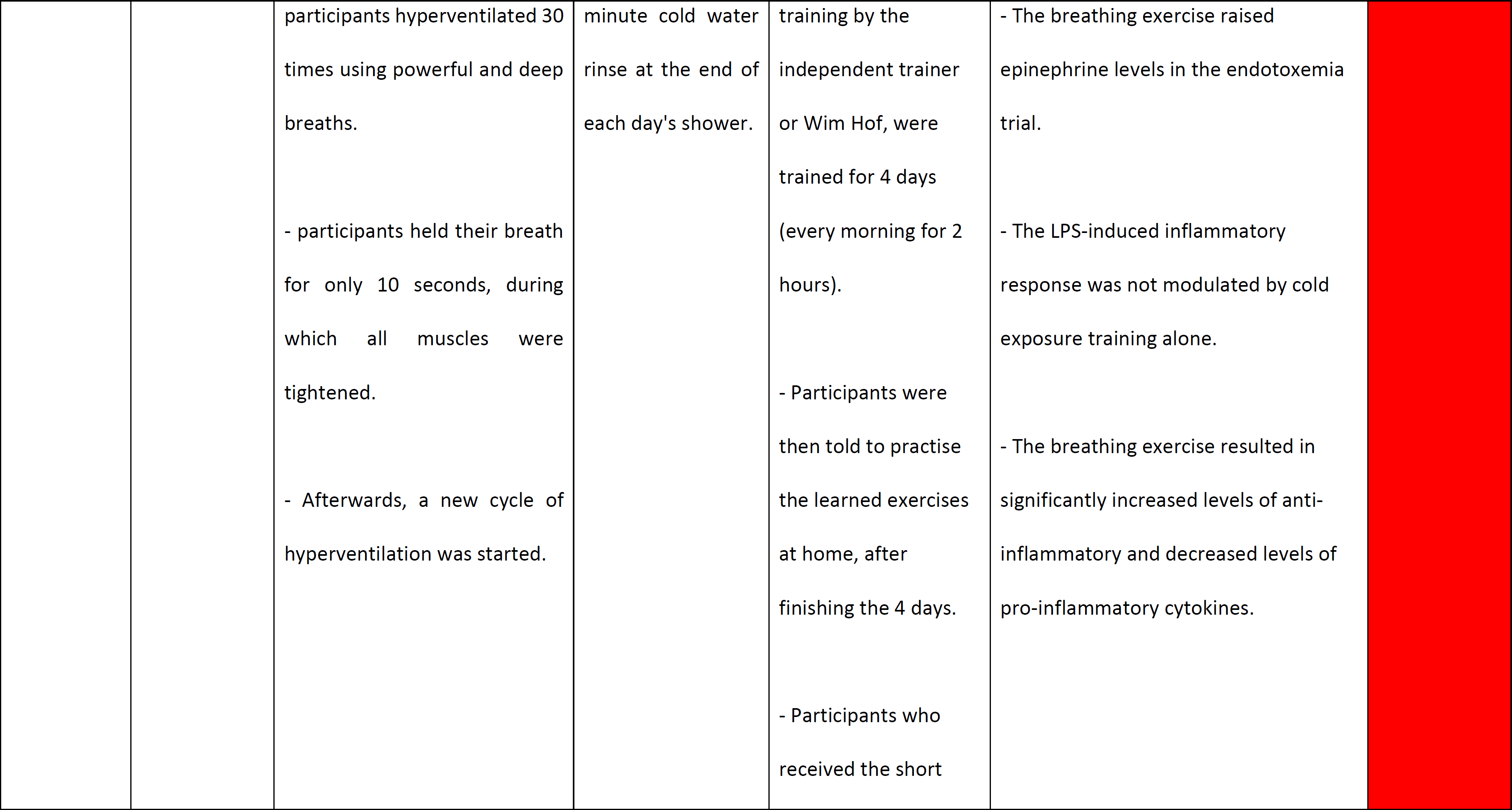
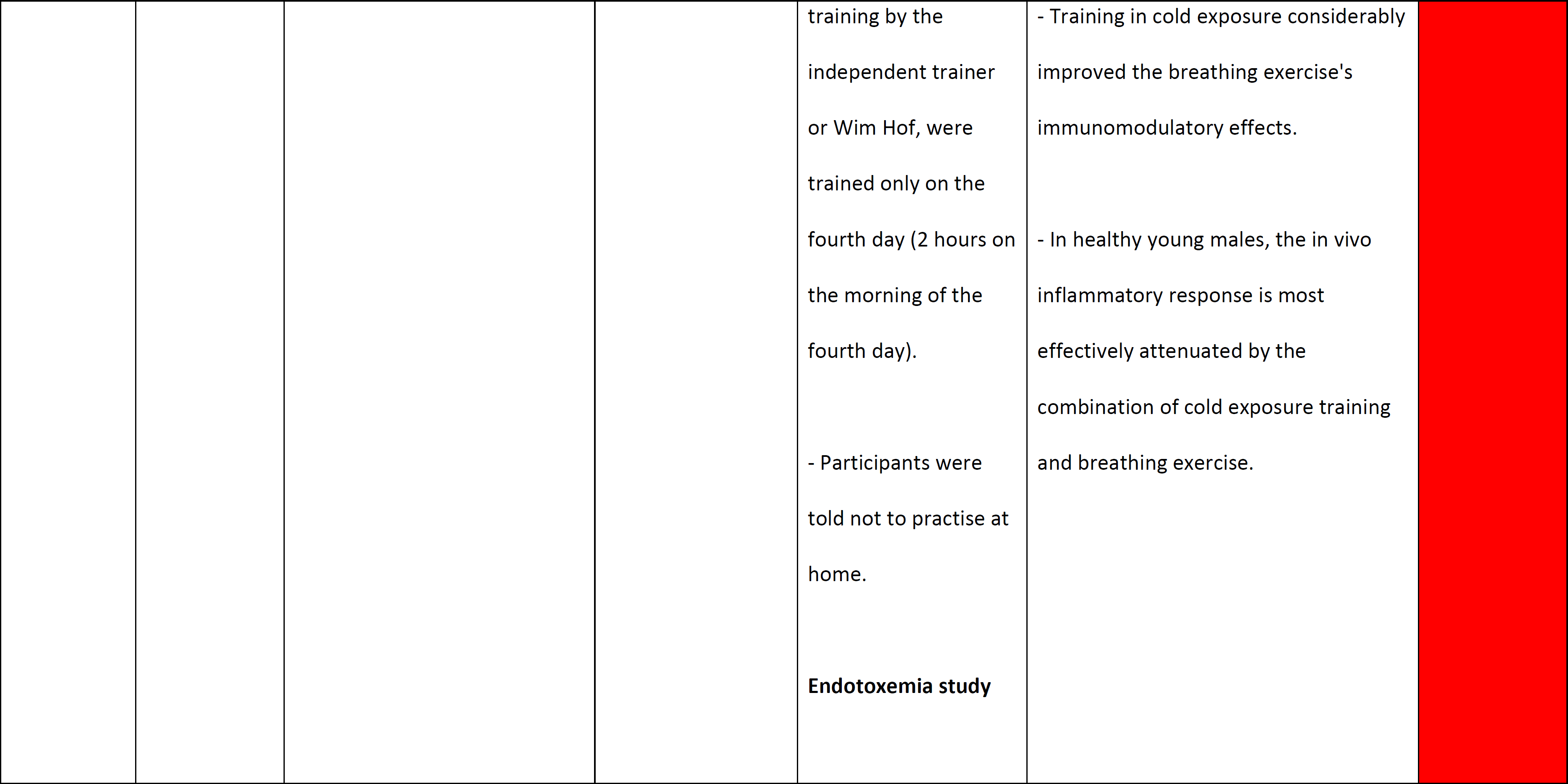
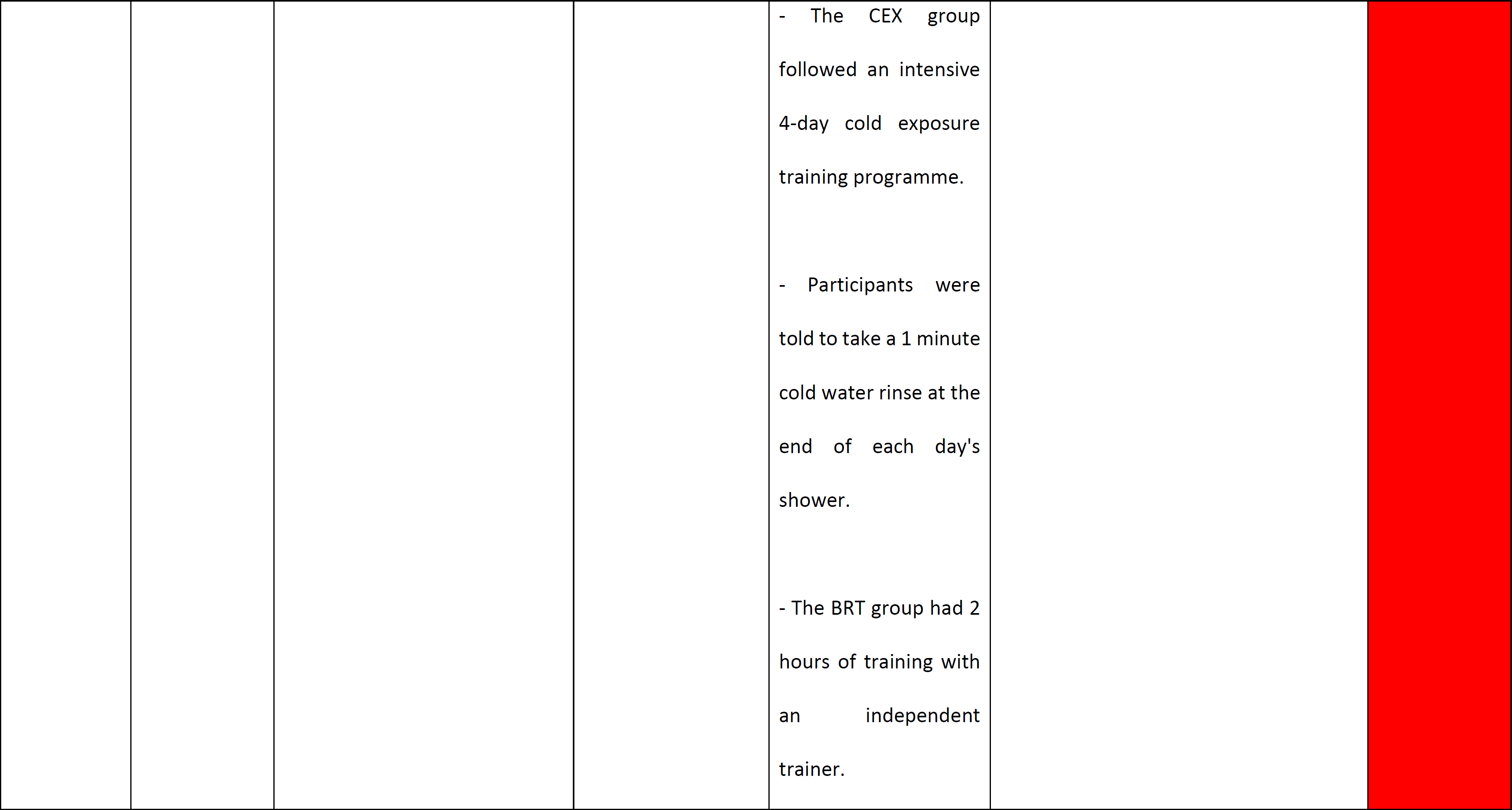
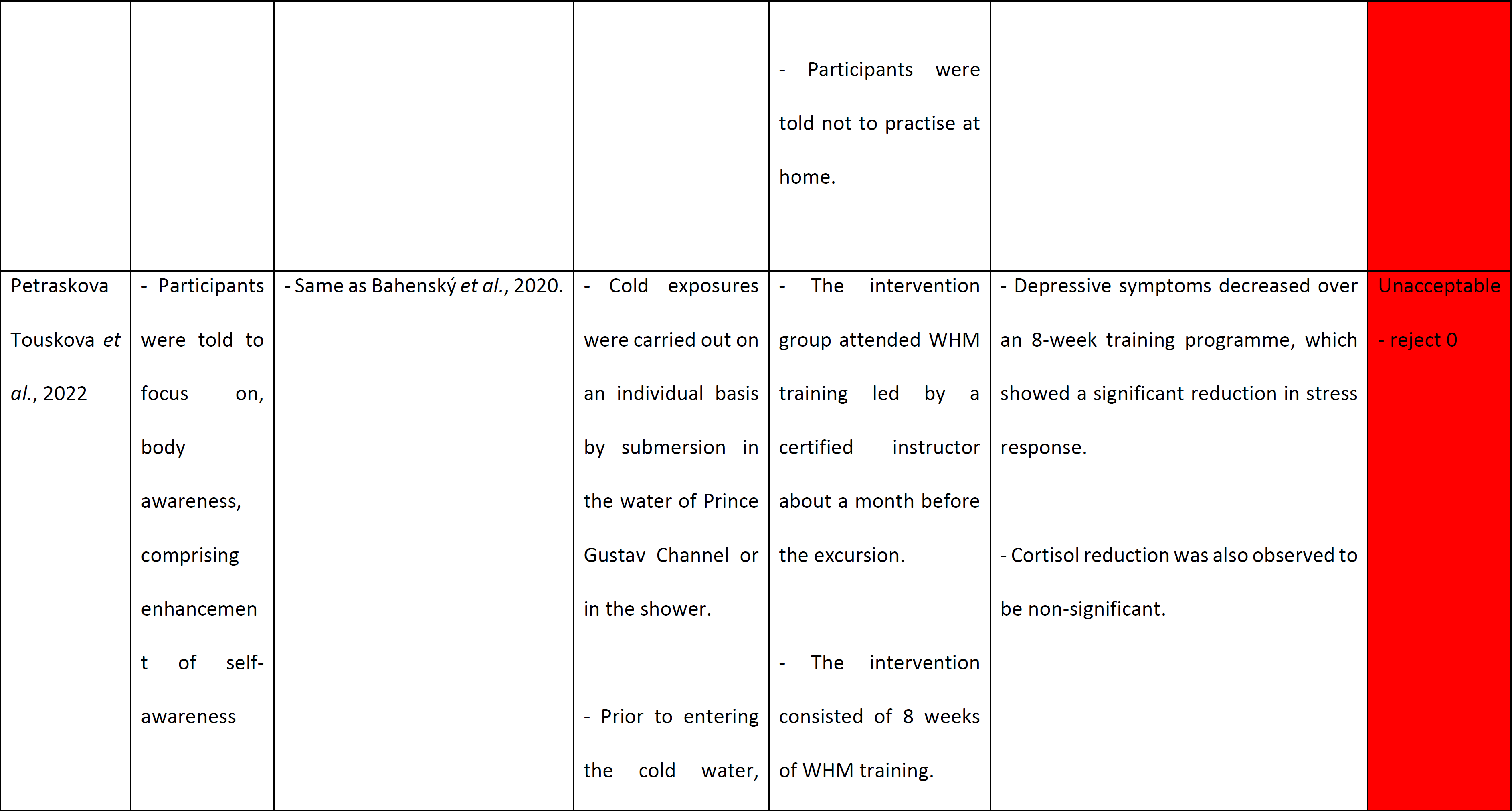
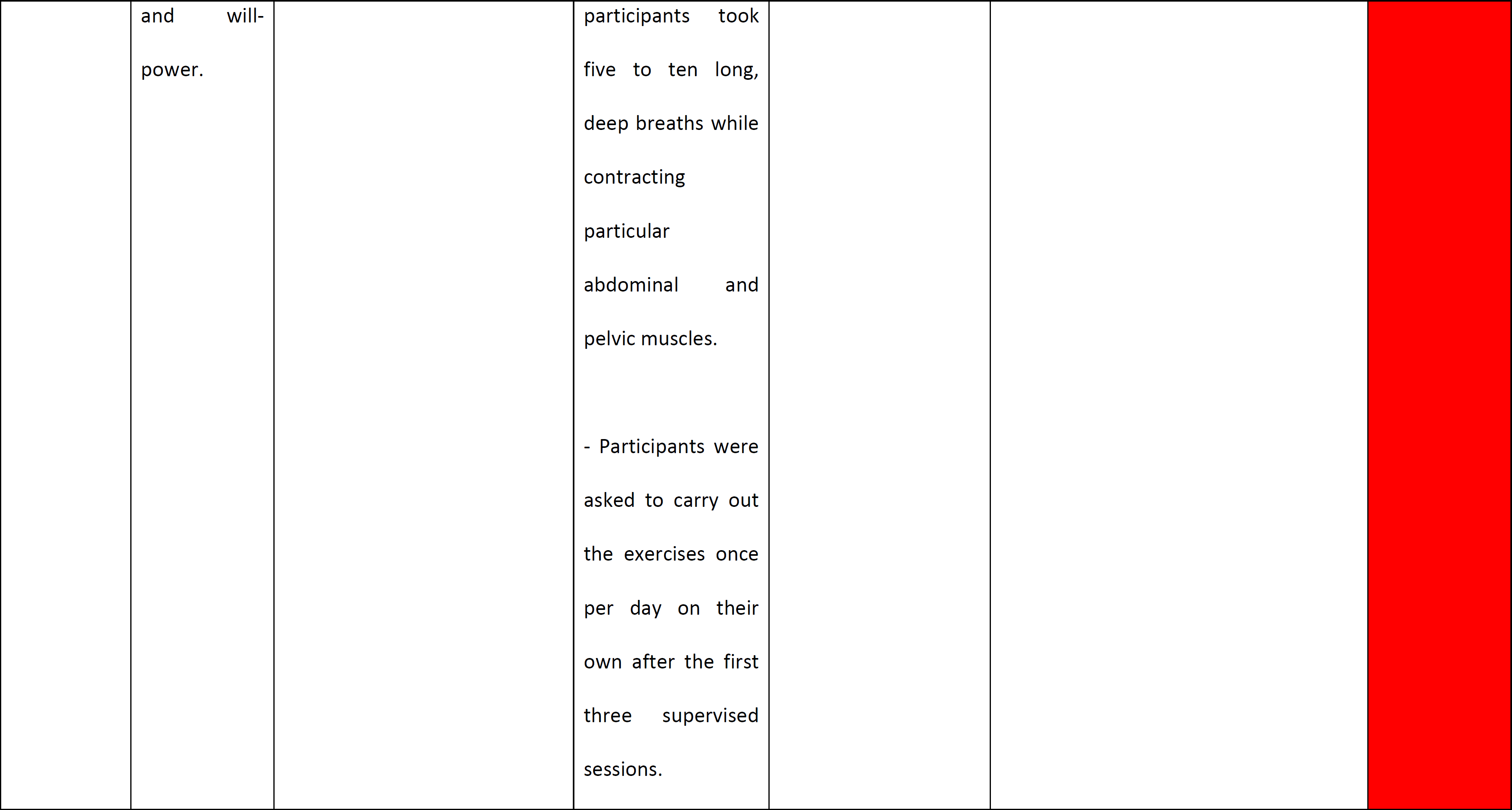
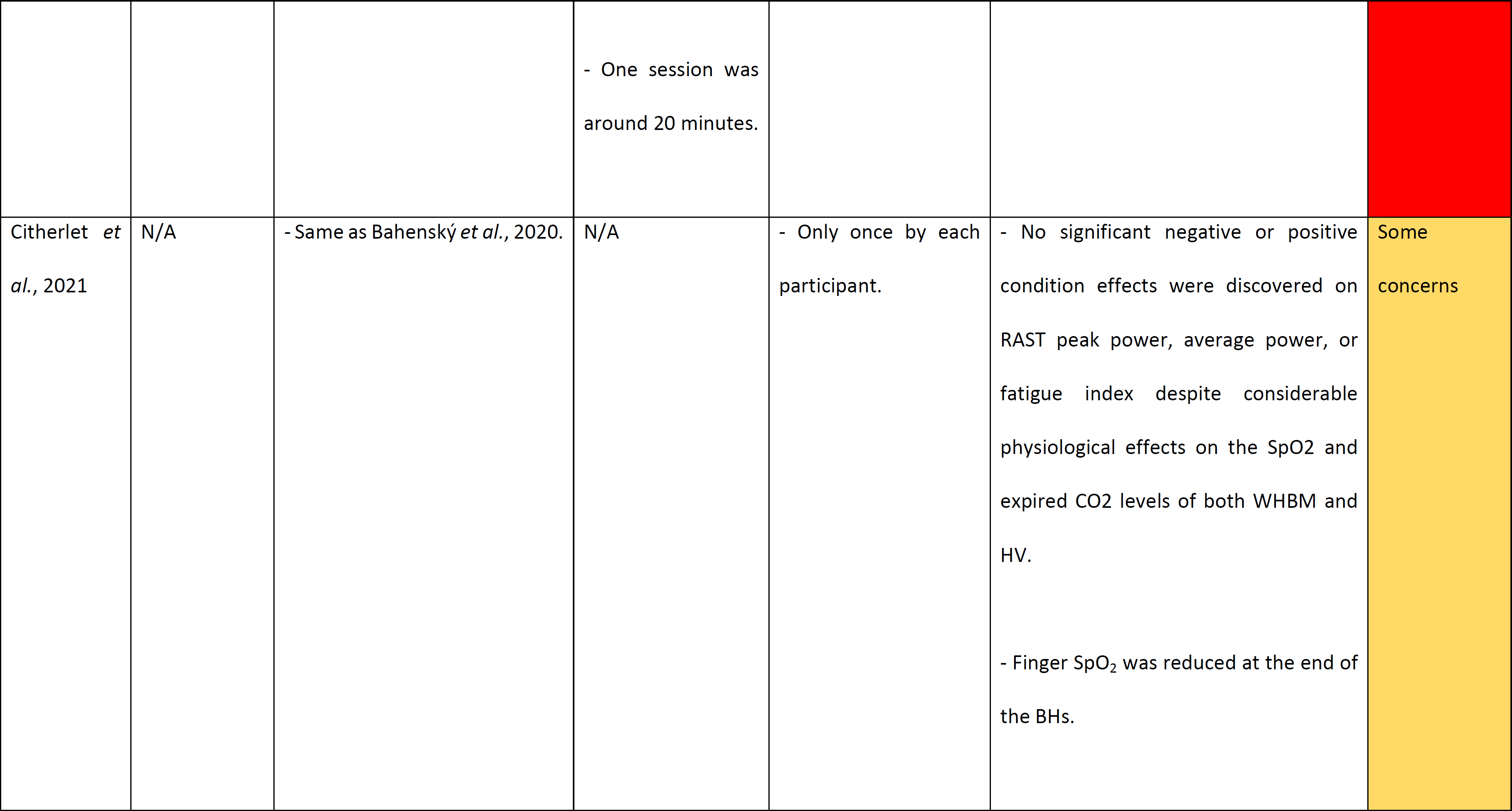
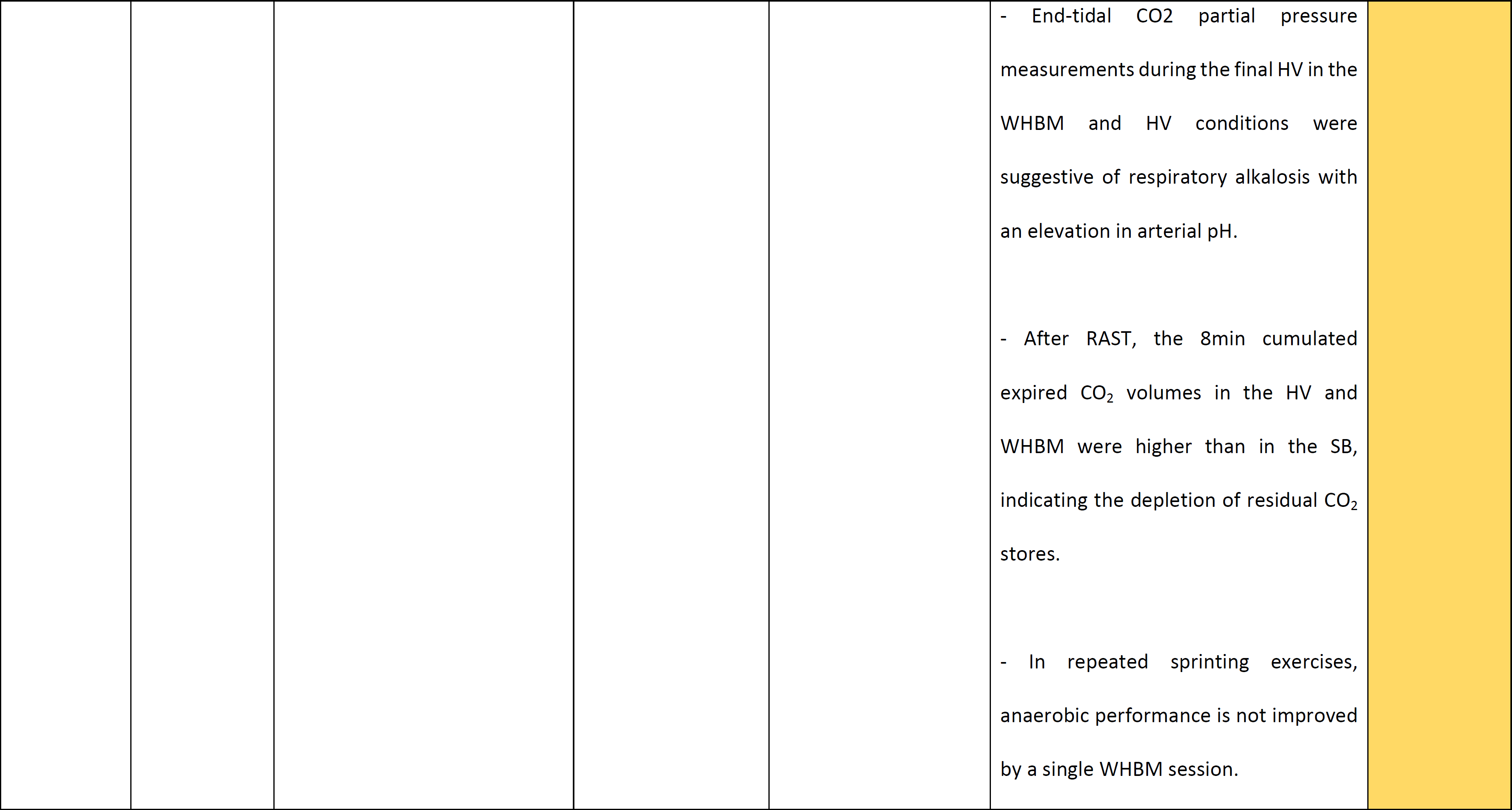
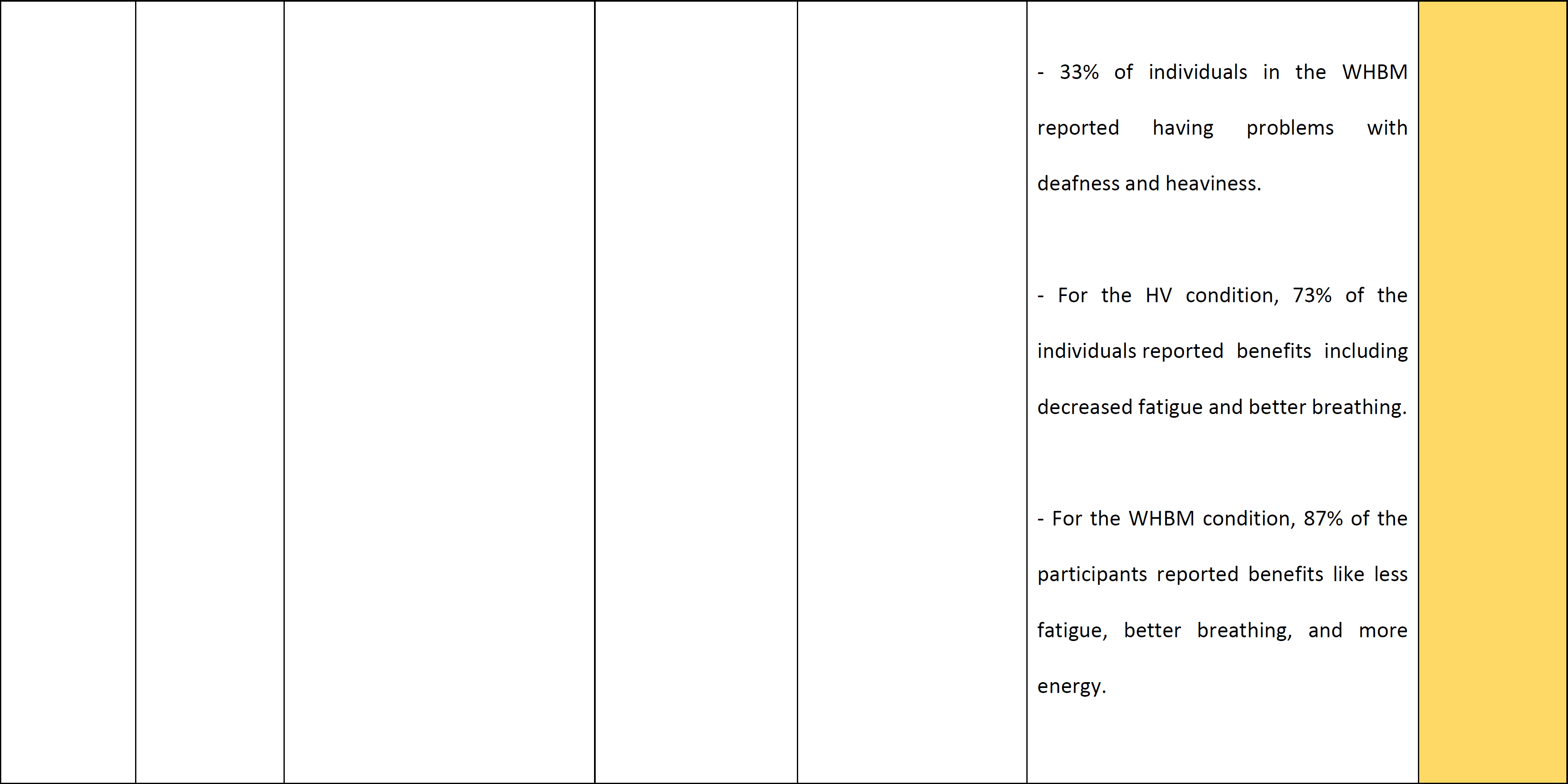
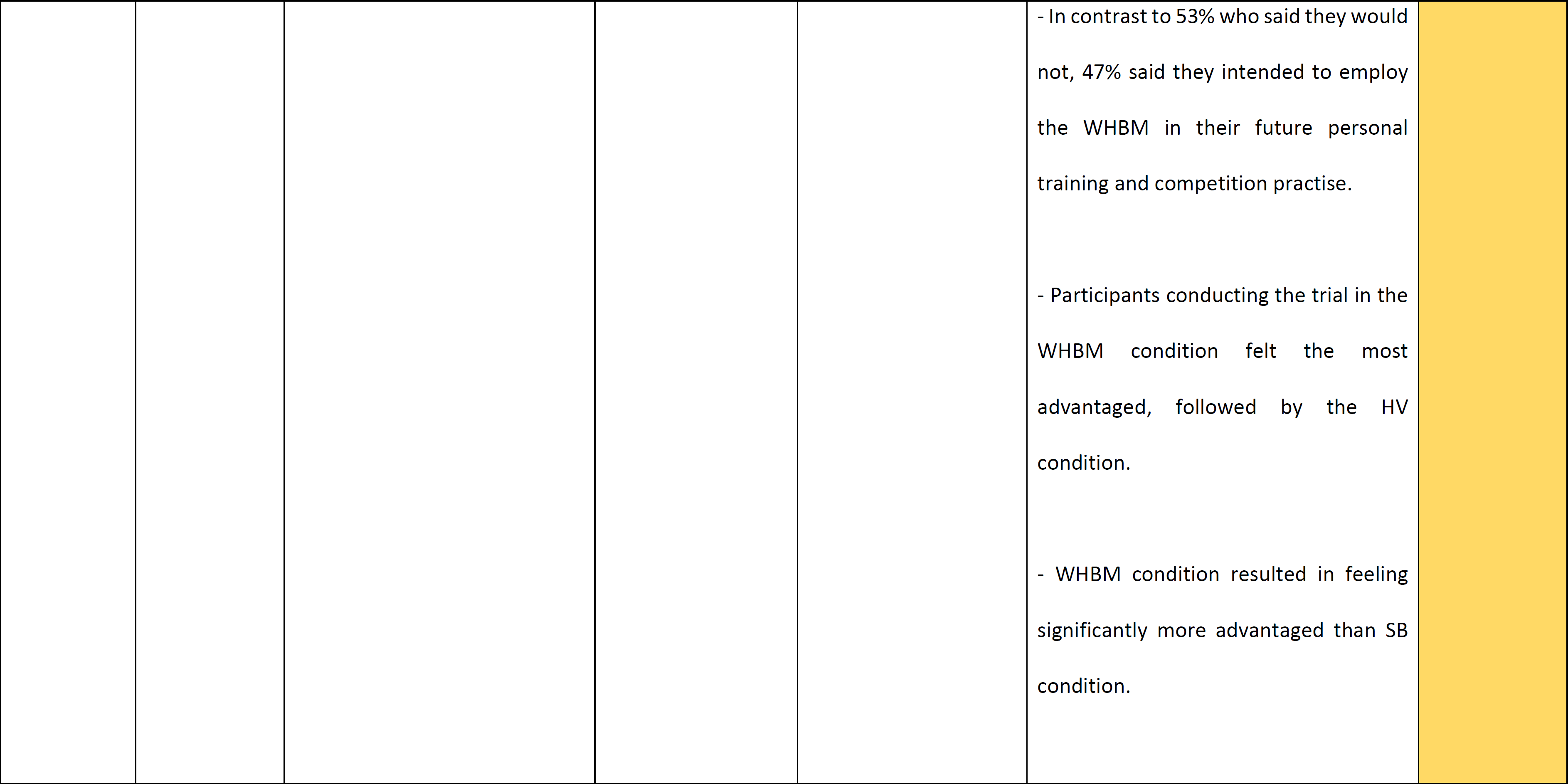
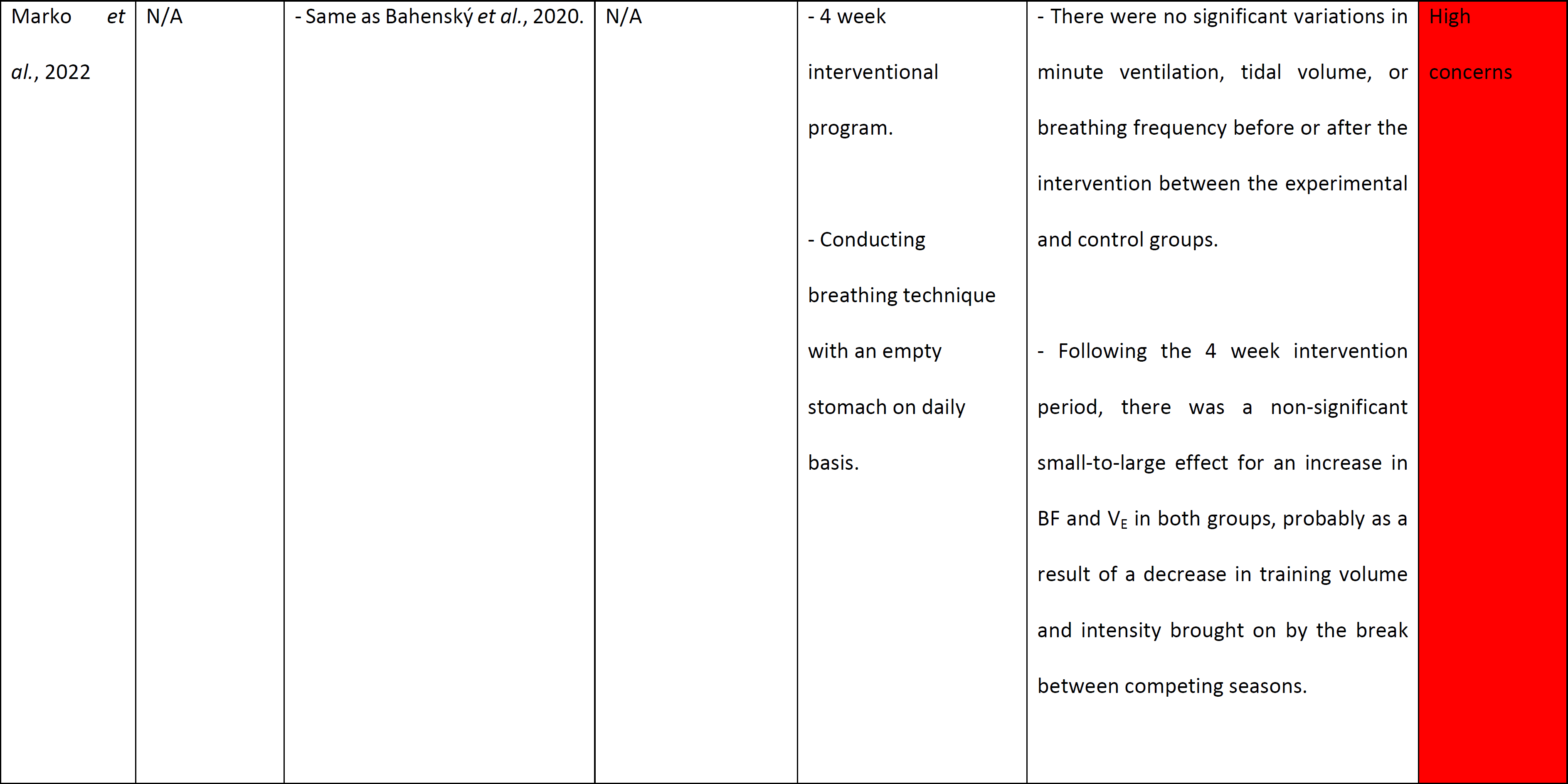
Summary of WHM description, key findings, and quality assessment.

### Stress response

Two trials investigated epinephrine levels during the endotoxemia experiment [10, 15] and one trial studied epinephrine levels during WHBM [15]. Baseline epinephrine levels in WHM-trained participants were significantly higher compared to non-trained participants (p=0.007). After starting WHBM, epinephrine levels increased further in this group and peaked just before endotoxemia administration and remained elevated until WHBM was stopped [10]. Additionally, in Zwaag *et al.*, 2022 endotoxemia study, participants had no variations in epinephrine levels throughout the experiment (p=0.48) in both participants exposed to cold (CEX) and participants not performing the WHM (CON). However, an increase in epinephrine levels began much earlier and was significantly greater in both the cold exposure with breathing exercise without retention (CBR) (p=0.01) and breathing exercise without retention (BRT) (p=0.04) compared to the CEX and CON groups [15]. Furthermore, in Zwaag *et al.*, 2022 breathing exercises study, both breathing exercises (WHBM with/without retention) raised epinephrine levels, which were unaffected by training duration (short vs extensive training: p=0.71) or the trainer (independent trainer vs Wim Hof: p=0.46). However, epinephrine was slightly more sustained in participants performing WHBM with retention compared to without retention (p=0.003) [15]. Moreover, Kox *et al.*, 2014 explored norepinephrine and dopamine levels during the endotoxemia trial and found them to be within the reference range throughout the experiment [10].

The associations of cortisol and melatonin with WHM were also explored. In Petraskova Touskova *et al.*, 2022 study, the group not practising WHM had higher, but statistically non-significant cortisol levels at the end of the expedition (p=0.327) compared to WHM-trained participants [12]. Whereas Kox *et al.*, 2014 found no variations in serum cortisol levels between the WHM-trained and non-trained groups before or throughout the period of WHM training; however, levels normalised faster in WHM-trained participants [10]. The difference in the results could be due to the different collection procedures for cortisol. Kox *et al.*, 2014 collected cortisol from blood while Petraskova Touskova *et al.*, 2022 collected hair cortisol. Furthermore, significantly lower levels of melatonin were found in the WHM compared to the non-trained group (p=0.03) [12].

Four trials examined HR recordings [6, 8, 10, 15]. Kox *et al.*, 2014 showed an increase in HR after starting WHBM which normalised faster in WHM-trained group compared to non-trained group [10]. Furthermore, Bahenský *et al.*, 2020 reported that HR tended to be lower after WHBM compared to no WHBM [6]. Citherlet *et al.*, 2021 also found HR significantly increased during hyperventilation (HV) and significantly decreased during breath hold in participants practising WHBM when compared to resting values [8]. Similarly, Zwaag *et al.*, 2022 observed a significant increase in HR in both CBR (p<0.001) and BRT (p<0.001) groups, during the first hyperventilation cycle, compared to the CON group. No changes in HR responses between CEX and CON groups (p=0.89) were found during the endotoxemia experiment. The HR measurements of BRT and CBR groups were comparable to those of CEX and CON groups when WHBM was stopped [15].

### Pro-inflammatory/anti-inflammatory responses

Two trials investigated cytokines levels during endotoxemia experiment [10, 15]. In Kox *et al.*, 2014 study, TNF-α, interleukin-6, and interleukin-8 levels were significantly lower in WHM-trained participants, whilst interleukin-10 levels were significantly higher (TNF-α, interleukin-6 and interleukin-8 levels were 53%, 57%, and 51% lower respectively; interleukin-10 levels were 194% higher) compared to non-trained group [10]. Similarly, Zwaag *et al.*, 2022 endotoxemia study, found TNF-α (p=0.03), interleukin-6 (p=0.03), and interleukin-8 (p<0.001) levels significantly lower, whereas interleukin-10 levels significantly higher (p=0.02) in CBR group than CON group. However, similar but less pronounced effects on cytokine levels were found when comparing BRT group to CON group for interleukin-6 (p=0.04) and interleukin-8 (p=0.02), but not for interleukin-10 (p=0.17). Additionally, CEX group did not show any significant changes in TNF-α (p=0.93), interleukin-6 (p=0.73), interleukin-8 (p=0.99), and interleukin-10 (p=0.44) levels compared to CON [15]. The differences seen might be due to the differences in WHBM. Kox *et al.*, 2014 performed WHBM with retention, while Zwaag *et al.*, 2022 endotoxemia study performed WHBM without retention.

Inflammation outcomes were also measured in participants with axial spondyloarthritis. During the 8-week WHM training, ESR, median BASDAI, ASDAS-CRP, and serum calprotectin decreased significantly (p=0.040, p=0.012, p=0.044, p=0.064 respectively) compared to participants not receiving WHM training. However, no statistical significance was found in hs-CRP between both groups (p=0.103) [7].

### Metabolites response

Zwaag *et al.*, 2020 showed that lactate and pyruvate play an important role in the anti-inflammatory response in WHM-trained participants. High pyruvate concentration, but not lactate concentration, increased endotoxemia-induced interleukin-10 production, and the combination of both metabolites resulted in an even more prominent and statistically robust increase. Although these two metabolites were highly intercorrelated (p<0.0001), they were not associated with the highly elevated epinephrine levels at any of the examined time points (p-values>0.15). Additionally, pyruvate reduced the production of interleukin-6. Lactate, pyruvate, and the two metabolites combined also tended to reduce endotoxemia-induced TNF-α production; however, significance was not achieved [16]. In Kox *et al.*, 2014 study, lactate levels were significantly higher but not to a clinically relevant level in WHM-trained compared to non-trained participants [10].

### Respiratory parameters

Three papers measured V_E_ [6, 8, 11] and two of them also measured V_T_ and BF [6, 11]. All three papers were investigating whether performing WHBM before an exercise will improve performance. Bahenský *et al.,* 2020 found the difference in mean values at each load to be significant for V_E_ [6] and Citherlet *et al.,* 2021 found a significant difference (p=0.039) between participants performing spontaneous breathing (SB) and WHBM at rest, but the mean difference was not statistically significant after performing an exercise (p>0.05) [8]. Additionally, Marko *et al.,* 2022 showed that at each load stage, V_E_ depicted statistical insignificance between WHBM and normal breathing (p=0.138, p=0.825, p=0.479, p=0.489) [11]. Furthermore, for V_T_ and BF measurements, Bahenský *et al.,* 2020 found the difference in mean values at each load to be significant (p<0.001, p<0.001) [6]; however, Marko *et al.,* 2022 showed no statistical significance between WHBM participants and normal breathing at any load stage (V_T_: p=0.630, p=0.377, p=0.688, p=0.087; BF: p=0.794, p=0.917, p=0.956, p=0.296) [11].

### Blood gas measurements

Two trials examined blood gas measurements. In Kox *et al.*, 2014 study, starting the WHBM resulted in an immediate and profound decrease in pCO_2_ and an increase in pH, but it was not stated if these changes were statistically significant. Furthermore, during WHBM training, the trained group’s oxygen saturation decreased significantly [10]. In contrast, Zwaag *et al.*, 2022 endotoxemia study, found pH and oxygen saturation levels significantly higher in the groups performing WHBM (BRT: p<0.01, p<0.001; CBR: p<0.01, p<0.001 respectively) when compared to CON. Whereas, pCO_2_ was significantly lower in the same groups (BRT: p<0.001; CBR: p<0.001) when compared to CON [15]. In Zwaag *et al.*, 2022 breathing exercises study, oxygen saturation significantly decreased at the end of each retention phase, only when WHBM is performed with retention (p<0.001) compared to without retention. While pH and pCO_2_ were similar in both WHBM exercises, except for a small but statistically significant difference at the final measurement time point (pH: p<0.001; pCO_2_: p<0.001) [15].

### Psychological response

One paper investigated the effect of optimism and neuroticism on WHM. It found that a higher level of expectancy and optimism helped significantly potentiate the effect of WHM while a decrease in neuroticism level was not found to significantly help. Participants in the training group were overall relatively optimistic and low in neuroticism. A higher level of optimism was associated with higher interleukin-10 levels (p<0.05) and epinephrine levels (p<0.01). Neuroticism was not found to be a significant predictor of endotoxin response. Participants’ expectations to overcome the endotoxemia experiment significantly increased from before WHM training to after the endotoxemia experiment (p=0.003) [13].

Another paper explored the quality of life of participants and found that SF-36 physical and mental component scores significantly increased over the WHM-training period (p=0.004, p=0.004 respectively) compared to the non-trained group. While EQ-5D and EQ-5D visual analogue scale did not experience a significant change between the WHM-trained and non-trained groups (EQ-5D: p=0.933, p=0.102; EQ-5D visual analogue scale: p=0.090, p=0.674) [7].

The last paper reported the results of the questionnaire and Borg RPE scale. Positive effects of increased energy, less fatigue, and improved breathing were reported by 87% of the participants. Almost half of the participants (47%) also said they would think about incorporating the WHBM into their own practices in the future. Additionally, when the experiment was performed using the WHBM, 66.7% of participants rated it the best method in terms of perceived performance compared to 13.3% for SB and 20% for HV. Likewise, 73.3% of participants judged the WHBM to be the best method to perform, while 53.3% judged SB as the worst. However, 33% of the participants using WHBM reported negative effects of deafness and heaviness [8]. Moreover, Borg RPE scale was found statistically significant in Bahenský *et al.*, 2020 (p<0.001) and Citherlet *et al.*, 2021 (WHBM compared to HV (p=0.008) and SB (p=0.017)), meaning that the intensity of a training session is less when WHM is being used [6, 8].

### Reporting of symptoms

Three papers reported experience of flu-like symptoms following endotoxemia administration. Kox *et al.*, 2014 reported lower flu-like and self-reported symptoms and faster recovery from fever in participants performing WHM [10]. Similarly, Middendorp *et al.*, 2016 showed that a higher expectation of the training’s effects was associated with lower flu-like clinical symptoms (p<0.01) compared to non-trained group [13]. Likewise, Zwaag *et al.*, 2022 endotoxemia study results showed that only CEX group had significantly lower flu-like symptoms (p=0.017) compared to CON group. All other groups had comparable peak symptom scores to CON group (BRT: (p=0.70); CBR: (p=0.21)). Additionally, when compared to the CON group, symptoms disappeared considerably faster in all three intervention groups (BRT: p<0.001; CEX: p=0.01; CBR: p=0.002) [15].

Two papers also reported depressive symptoms. Petraskova Touskova *et al.*, 2022 found that at the end of the expedition, depressive symptoms were significantly lower in the WHM-trained group compared to non-trained group (p=0.03), while TSC-40 scores, which measures stess-related symptoms, were higher in the non-trained group, but not significantly [12]. Additionally, Buijze *et al.*, 2019 found no significant effect on depressive symptoms between WHM-trained and non-trained groups [7].

The final paper reported AEs in axial spondyloarthritis participants [7]. Buijze *et al.*, 2019 found no significant differences in AEs between WHM-trained and non-trained participants, with just one serious AE (hypertensive crisis) occurring 8 weeks after the completion of the intervention and was judged unrelated [7].

## Discussion

This is the first systematic review conducted on WHM. The findings of this review suggest that WHM may affect the reduction of inflammation in healthy and non-healthy participants and that the WHBM does not enhance the performance of an exercise. However despite the statistical significance observed in some studies, it must be noted that the quality of the studies is very low, meaning that all the results must be interpreted with caution. Additionally, the low sample size (15-48 individuals per study) and large proportion of males in the studies (86.4%) make the results non-generalizable to the public. Consideration should also be given that participants might experience the placebo effect, where improvements in patients’ symptoms are due to their participation in the therapeutic encounter, with its rituals, symbols, and interactions [26]. For example, *Middendorp et al.,* (2016) found that a higher level of expectancy and optimism helped significantly potentiate the effect of WHM [13]. However, Zwaag *et al.,* (2022), who queried the so-called ‘guru effect’ [27] and whether it is necessary to be trained by the creator of the intervention to influences symptomatology [15], found that observed physiological and immunological effects are independent from the individual who provides the WHM treatment intervention.

Out of all the categories, WHM appears to have the most benefit in the stress and pro-inflammatory/anti-inflammatory response categories. Inflammation, especially when chronic, can cause severe complications such as cardiovascular diseases, cancer, diabetes, rheumatoid arthritis, asthma, chronic obstructive pulmonary disease, alzheimer, chronic kidney disease, and inflammatory bowel disease, therefore, reducing inflammation is beneficial [28]. The WHM reduces inflammation using a different mechanism of action (MOA) than other anti-inflammatory interventions. The closest MOA to that of WHM is corticosteroids’ MOA as they both increase interleukin-10 production. WHM increases epinephrine, causing an increase in interleukin-10 which leads to a reduction in pro-inflammatory cytokines [10, 15]. Whereas corticosteroids activate anti-inflammatory genes, including interleukin-10 gene, and inactivate multiple inflammatory genes by inhibiting histone acetyltransferase and recruiting histone deacetylase-2 activity to the inflammatory gene transcriptional complex [29]. The findings of this review suggest that WHM may provide some benefits in healthy and non-healthy people as it was suggested to be safe and might decrease inflammation, unlike corticosteroids, which are only given to non-healthy people due to having many side effects.

The least benefited category of the WHM was the respiratory parameters. Although WHBM was found to not enhance exercise performance, it could have other potential applications. For example, a published letter suggests that WHBM may be helpful in preventing and reversing symptoms of acute mountain sickness (AMS) [30]. Wim Hof and a combination of healthy and non-healthy participants climbed a mountain using the WHBM in 2 days instead of the usual 4-7 days and none of the participants suffered from severe AMS. These findings suggest that a trial on this topic may be of importance, especially for the rescue teams that must ascend quickly [30]. However, although this letter was published in a journal, it was not peer-reviewed hence the results should be interpreted with caution.

There are three pillars to WHM: cold exposure, WHBM, and meditation. The latter is a foundation of the other two pillars, thus there were no studies testing it by itself. In this review, cold exposure alone was suggested to have an insignificant effect on epinephrine and cytokine levels [15]. Similarly, the wider literature agrees with this finding. For example, cold water immersion (CWI) did activate the immune system and alter systematic inflammation but not significantly [31, 32, 33, 34]. Only one study found CWI helpful in reducing inflammation in rock climbers by reducing vessel permeability toward the site of inflammation [35]. Furthermore, in this review, WHBM alone was claimed to significantly increase epinephrine levels, regardless of the duration of training or trainer, and that increase was more prolonged in participants performing WHBM with retention [15]. On the other hand, cytokine levels were only measured in participants performing WHBM without retention. There were no trials comparing WHBM with retention alone on cytokine levels. When WHBM was performed without retention, pro-inflammatory cytokines significantly decreased, but interleukin-10 levels were not increased significantly [15]. This might be because hypoxia improves interleukin-10 release and reduces the pro-inflammatory response via improved adenosine release [36]. Thus, it would be interesting for future trials to investigate the effects of WHBM with retention alone on cytokine levels. Nevertheless, when cold exposure was combined with WHBM, regardless of whether it was with retention or without, statistical significance was achieved on epinephrine and cytokine levels [10, 15]. Therefore, it is probably best to perform all the pillars of WHM to achieve a significant improvement in immunomodulatory effects. Future trials should investigate whether there is a significant difference between performing WHBM with retention and cold exposure compared to WHBM without retention and cold exposure on epinephrine and cytokine levels.

This review suffers from several limitations. Firstly, the outcomes were very heterogeneous as many unrelated outcomes are included. This review could have focused on synthesising specific outcomes such as only focusing on outcomes related to stress and inflammation. Secondly, all the trials had a very high risk of bias. This was due to the lack of a prior published protocol, small sample size, and complexity of blinding the participants and outcome assessors to the intervention. Thirdly, some outcomes such as psychological outcomes were difficult to measure as they are subjective measures usually assessed using a questionnaire. Since the participants were not blinded, it was very difficult to ensure that the answers were honest and valid to the experience. Lastly, the sample size was very small, sometimes affecting the results as any error in measurement or a loss of follow-up can significantly shift the trial results. More evidence needs to be synthesised about WHM before being recommended to the public. Even though it is very difficult to prove it scientifically as conducting a double-blinded RCT, which is the gold standard, is impossible because participants cannot be blinded to the WHM. It does appear to offer benefits in attenuating inflammation with minimal serious adverse events and good positive effects of increased energy, less fatigue, and improved breathing [7, 8, 10, 15]. Thus, WHM can probably be used by lifestyle medicine to decrease inflammation in people suffering from inflammatory disorders. However, to be able to recommend WHM in an unwell population, future trials should publish a protocol outlining their experiment before starting the trial, increase the sample size, and make sure to account for loss to follow-up. Additionally, researchers should make sure to blind outcome assessors, and ensure that the outcomes assessed are objective and not subjective.

## Conclusion

Considering all the studies, the WHM may produce promising immunomodulatory effects but more research of higher quality is needed to substantiate this finding. The combination of cold exposure and WHBM appeared to most effectively reduce the inflammatory response. Hence, all the pillars of the WHM are important to extract the benefits. The focus of future studies should move away from investigating the use of WHBM to enhance exercise performance and towards exploring the benefits of WHM in non-healthy participants. WHM could be further studied in preventing or treating diseases, such as inflammatory disorders. Studies about WHM have not yet investigated all the beneficial claims the WHM states to have. Future studies may provide valuable insights about WHM as there is still much to explore.

## Data Availability

All relevant data are within the manuscript and its Supporting Information files.

## Acknowledgements

None

## References

1. Hof W. Who is ‘The Iceman’ Wim Hof? 2022 [Available from: https://www.wimhofmethod.com/iceman-wim-hof.

2. Hof W. Wim Hof Method breathing 2022 [Available from: https://www.wimhofmethod.com/breathing-exercises.

3. Hof W. Cold therapy 2022 [Available from: https://www.wimhofmethod.com/cold-therapy.

4. Hof W. How to increase willpower 2022 [Available from: https://www.wimhofmethod.com/how-to-increase-willpower.

5. Muzik O, Reilly KT, Diwadkar VA. “Brain over body”–A study on the willful regulation of autonomic function during cold exposure. NeuroImage. 2018;172:632–41.

6. Bahenský P, Marko D, Grosicki GJ, Malátová R. Warm-up breathing exercises accelerate VO2 kinetics and reduce subjective strain during incremental cycling exercise in adolescents. Journal of Physical Education and Sport. 2020;20(online ISSN: 2247 - 806X; p-ISSN: 2247 – 8051; ISSN - L = 2247 - 8051).

7. Buijze GA, De Jong HMY, Kox M, van de Sande MG, Van Schaardenburg D, Van Vugt RM, et al. An add-on training program involving breathing exercises, cold exposure, and meditation attenuates inflammation and disease activity in axial spondyloarthritis - A proof of concept trial. PloS one. 2019;14(12):e0225749.

8. Citherlet T, Crettaz von Roten F, Kayser B, Guex K. Acute Effects of the Wim Hof Breathing Method on Repeated Sprint Ability: A Pilot Study. Frontiers in sports and active living. 2021;3:700757.

9. Kox M, Stoffels M, Smeekens SP, van Alfen N, Gomes M, Eijsvogels TM, et al. The influence of concentration/meditation on autonomic nervous system activity and the innate immune response: a case study. Psychosom Med. 2012;74(5):489–94.

10. Kox M, van Eijk LT, Zwaag J, van den Wildenberg J, Sweep F, van der Hoeven JG, et al. Voluntary activation of the sympathetic nervous system and attenuation of the innate immune response in humans. PROCEEDINGS OF THE NATIONAL ACADEMY OF SCIENCES OF THE UNITED STATES OF AMERICA. 2014;111(20):7379–84.

11. Marko D, Bahensky P, Bunc V, Grosicki GJ, Vondrasek JD. Does Wim Hof Method Improve Breathing Economy during Exercise? Journal of clinical medicine. 2022;11(8).

12. Petraskova Touskova T, Bob P, Bares Z, Vanickova Z, Nyvlt D, Raboch J. A novel Wim Hof psychophysiological training program to reduce stress responses during an Antarctic expedition. The Journal of international medical research. 2022;50(4):3000605221089883.

13. Middendorp Hv, Kox M, Pickkers P, Evers AWM. The role of outcome expectancies for a training program consisting of meditation, breathing exercises, and cold exposure on the response to endotoxin administration: a proof-of-principle study. CLINICAL RHEUMATOLOGY. 2016;35(4):1081–5.

14. Vosselman MJ, Vijgen GHEJ, Kingma BRM, Brans B, van Marken Lichtenbelt WD. Frequent Extreme Cold Exposure and Brown Fat and Cold-Induced Thermogenesis: A Study in a Monozygotic Twin. PLOS ONE. 2014;9(7):e101653.

15. Zwaag J, Naaktgeboren R, van Herwaarden AE, Pickkers P, Kox M. The Effects of Cold Exposure Training and a Breathing Exercise on the Inflammatory Response in Humans: A Pilot Study. Psychosomatic medicine. 2022;84(4):457–67.

16. Zwaag J, ter Horst R, Blazenovic I, Stoessel D, Ratter J, Worseck JM, et al. Involvement of Lactate and Pyruvate in the Anti-Inflammatory Effects Exerted by Voluntary Activation of the Sympathetic Nervous System. METABOLITES. 2020;10(4).

17. Hof W. WIm Hof Method benefits 2022 [Available from: https://www.wimhofmethod.com/benefits.

18. Rippe JM. Lifestyle Medicine: The Health Promoting Power of Daily Habits and Practices. Am J Lifestyle Med. 2018;12(6):499–512.

19. Rojek C. Celebrity and religion. Stardom and celebrity: A reader. 2007:171–80.

20. Baker SA, Rojek C. The Belle Gibson scandal: The rise of lifestyle gurus as micro-celebrities in low-trust societies. Journal of Sociology. 2020;56(3):388–404.

21. Nunan D, Blane DN, McCartney M. Exemplary medical care or Trojan horse? An analysis of the ‘lifestyle medicine’ movement. British Journal of General Practice. 2021;71(706):229.

22. Page MJ, McKenzie JE, Bossuyt PM, Boutron I, Hoffmann TC, Mulrow CD, et al. The PRISMA 2020 statement: an updated guideline for reporting systematic reviews. BMJ. 2021;372:n71.

23. Kozhevnikov M, Elliott J, Shephard J, Gramann K. Neurocognitive and Somatic Components of Temperature Increases during g-Tummo Meditation: Legend and Reality. PLOS ONE. 2013;8(3):e58244.

24. Higgins JP, Savović J, Page MJ, Elbers RG, Sterne JA. Cochrane Handbook for Systematic Reviews of Interventions. Chapter 8: Assessing risk of bias in a randomized trial2022.

25. Healthcare Improvement Scotland. Checklists 2021 [Available from: https://www.sign.ac.uk/what-we-do/methodology/checklists/.

26. Kaptchuk TJ, Miller FG. Placebo Effects in Medicine. New England Journal of Medicine. 2015;373(1):8–9.

27. Sperber D. The guru effect. Review of Philosophy and Psychology. 2010;1:583–92.

28. Pahwa R, Goyal A, Jialal I. Chronic Inflammation StatPearls2022 [Available from: https://www.ncbi.nlm.nih.gov/books/NBK493173/?report=classic.

29. Barnes PJ. How corticosteroids control inflammation: Quintiles Prize Lecture 2005. Br J Pharmacol. 2006;148(3):245–54.

30. Buijze GA, Hopman MT. Controlled Hyperventilation After Training May Accelerate Altitude Acclimatization. Wilderness & Environmental Medicine. 2014;25(4):484–6.

31. Peake JM, Roberts LA, Figueiredo VC, Egner I, Krog S, Aas SN, et al. The effects of cold water immersion and active recovery on inflammation and cell stress responses in human skeletal muscle after resistance exercise. J Physiol. 2017;595(3):695–711.

32. Siqueira AF, Vieira A, Bottaro M, Ferreira-Júnior JB, Nóbrega OdT, de Souza VC, et al. Multiple Cold-Water Immersions Attenuate Muscle Damage but not Alter Systemic Inflammation and Muscle Function Recovery: A Parallel Randomized Controlled Trial. Scientific Reports. 2018;8(1):10961.

33. Janský L, Pospísilová D, Honzová S, Ulicný B, Srámek P, Zeman V, et al. Immune system of cold-exposed and cold-adapted humans. Eur J Appl Physiol Occup Physiol. 1996;72(5-6):445–50.

34. Allan R, Mawhinney C. Is the ice bath finally melting? Cold water immersion is no greater than active recovery upon local and systemic inflammatory cellular stress in humans. J Physiol. 2017;595(6):1857-8.

35. Heyman E, De Geus BAS, Mertens I, Meeusen R. Effects of Four Recovery Methods on Repeated Maximal Rock Climbing Performance. Medicine & Science in Sports & Exercise. 2009;41(6).

36. Kiers D, Wielockx B, Peters E, van Eijk LT, Gerretsen J, John A, et al. Short-Term Hypoxia Dampens Inflammation in vivo via Enhanced Adenosine Release and Adenosine 2B Receptor Stimulation. EBioMedicine. 2018;33:144–56.

